# Pathological expansion of gut microbiome-associated *Enterococcus* in advanced cirrhosis corresponds with multilevel perturbations of the gut-liver-immune axis

**DOI:** 10.1101/2025.02.13.25322003

**Authors:** Matthew J Dalby, Raymond Kiu, Marilena Stamouli, Betsy Arefaine, Lex E X Leong, Jack Hales, Christine Bernsmeier, Arjuna Singanayagam, Sidsel Støy, Maria-Emanuela Maxan, Merianne Mohamad, Ane Zamalloa, Matt Lewis, Royce P. Vincent, I. Jane Cox, Adrien Le Guennec, Roger Williams, Lindsey A Edwards, Shilpa Chokshi, Mark Thursz, Naiara Beraza, Charalambos G Antoniades, Debbie L Shawcross, Geraint B Rogers, Julia A Wendon, Kenneth D Bruce, Lindsay J Hall, Vishal C Patel

## Abstract

**Background:** Chronic liver disease (CLD) is a progressive condition that can advance to cirrhosis and acute-on-chronic liver failure (ACLF), a syndrome characterised by multi-organ dysfunction, critical illness, and high mortality. ACLF is driven by systemic inflammation, often without overt infection, suggesting alternative immune activation pathways, including microbial translocation. While intestinal perturbations, bacterial translocation, and immune dysfunction are hallmarks of ACLF, the specific microbial contributors remain unclear.

**Objective:** To investigate relationships between gut microbiome alterations, systemic inflammation, and clinical markers of disease severity across the cirrhosis spectrum.

**Design:** This cross-sectional, prospective study analysed faecal microbiota, plasma bile acids, urinary and plasma metabolites, gut and systemic inflammation and translocation markers, and monocyte dysfunction, in a well-phenotyped cohort of ACLF patients, decompensated and stable cirrhosis, compared to healthy individuals.

**Results:** Advanced stages of cirrhosis exhibited higher *Enterococcus* abundance, correlating with systemic inflammation, particularly in ACLF patients. Anaerobic commensal genera (*Roseburia*, *Ruminococcus*, and *Faecalibacterium*) were significantly lower. Lower urinary hippurate and trimethylamine N-oxide (TMAO) levels, linked to reduced microbial metabolism, paralleled these microbiome changes. Systemic inflammatory markers suggested parallel gut barrier dysfunction and microbial translocation in advanced cirrhosis.

**Conclusion:** Intestinal *Enterococcus* abundance, in advanced cirrhosis with greater antibiotic exposure, is a potential driver of gut barrier inflammation and dysfunction, and systemic immune activation. Further research into tailored microbiome-targeted therapies, including prebiotics, probiotics, phages and focused antibiotic use may prevent *Enterococcus* dominance, restore gut-liver axis homeostasis, and mitigate disease progression in cirrhosis.

Graphical Abstract

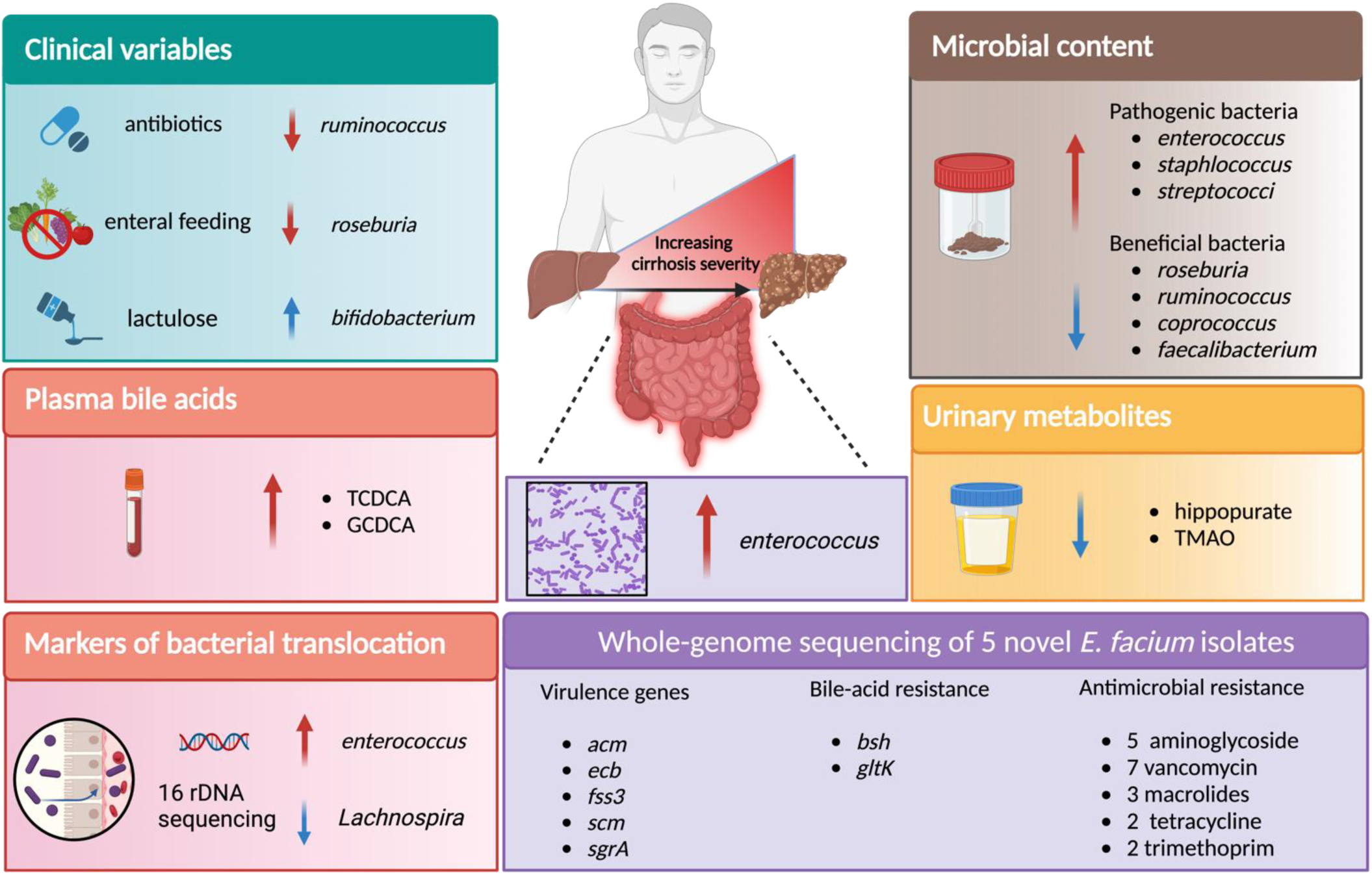

**Key messages:** *What is already known on this topic:*

- CLD can progress to ACLF, a life-threatening syndrome with high mortality rates characterised by systemic inflammation, impaired phagocytosis and immune dysfunction with associated multiple organ failure.
- The gut microbiome’s role in underpinning the gut-liver axis is recognised, with microbial perturbations and gut barrier dysfunction associated with CLD, yet the exact microbiome changes and their link to inflammatory pathways remain unclear.

*What this study adds:*

- This study reveals that *Enterococcus* significantly increases with CLD severity and antibiotic use and correlates with markers of systemic inflammation and gut barrier dysfunction.
- Reductions in obligate anaerobe commensal bacteria, such as *Roseburia* and *Faecalibacterium*, are observed, alongside depressed levels of key microbial metabolites, including hippurate and TMAO, across CLD stages.
- These changes suggest that *Enterococcus* predominance and gut microbiome perturbations driven by factors including disease severity and antibiotic use may actively drive inflammation and impaired monocyte function in CLD.

*How this study might affect research, practice, or policy:*

- Findings indicate that *Enterococcus* and wider gut microbiome perturbations provide a rationale for targeting as therapeutic strategies to modulate the gut-liver axis in CLD.
- This highlights the potential of microbiome-focused interventions, such as faecal microbial transplantation, probiotics, prebiotics or phages, to improve gut health, reduce gut and systemic inflammation, and ultimately mitigate cirrhosis-related complications.
- Future research and development of targeted microbiome therapeutics and rapid infection diagnostics may positively influence CLD management, including a focus on antimicrobial stewardship and informing clinical guidelines whilst improving patient outcomes.

## 1. Introduction

Chronic liver disease (CLD) is a progressive condition, resulting from several different aetiologies such as alcohol, viral hepatitis, and obesity that can culminate in cirrhosis and eventual liver failure (Huang et al., 2023). An important aspect of CLD is the development of acute-on-chronic liver failure (ACLF), a distinct syndrome characterised by multi-organ dysfunction, worsening liver function, and liver failure, with high mortality rates (Katoonizadeh et al., 2010; Moreau et al., 2013; Shah et al., 2024). The transition to decompensated cirrhosis, preceding ACLF, can lack identifiable triggers, confounding therapeutic options, exacerbating morbidity, and reducing transplant-free survival (D’Amico et al., 2022; Trebicka et al., 2021).

The gut microbiota is a key component of the ‘gut-liver axis’ but its interplay with the disease mechanisms is still not fully understood in CLD (Albillos et al., 2020; Hsu and Schnabl, 2023). While bacterial infections can act as a trigger of progression to ACLF, many cases (60-70%) display systemic inflammation without identifiable infection, with 50% of patients with ‘sepsis’ culture negative, suggesting alternative inflammatory pathways and limitations in infection diagnosis methods (Hernaez et al., 2017; Toledo et al., 1993). This ‘sterile inflammation’ in CLD patients may be driven by damage-associated molecular patterns (DAMPs) released from stressed or dying hepatocytes particularly in those without clear triggers such as active alcohol use or culture-confirmed bacterial infections (Arroyo et al., 2015). Additionally, translocation of viable bacteria or microbial components (i.e., microbial-associated molecular patterns [MAMPs]) *via* a dysfunctional intestinal barrier, may contribute to systemic inflammation, precipitating cirrhosis-related complications. Increased markers of gut permeability and inflammation in CLD patients further support the link between microbial translocation and inflammatory responses (Alexopoulou et al., 2017; Riva et al., 2020).

The gut microbiota potentially plays a dual role in CLD, serving as a reservoir for pathogens and MAMPs, while also simultaneously maintaining gut homeostasis through its resident commensal bacteria (Adak and Khan, 2019; Caballero-Flores et al., 2023). These commensal anaerobic microbes are critical for intestinal epithelial integrity, pathobiont exclusion (and preventing overgrowth), and metabolite production (e.g. short-chain fatty acids [SCFAs]) essential for host health (Ghosh et al., 2021). In CLD, however, gut microbiota disruptions are frequently observed, associated with reduced intestinal barrier integrity, leading to disrupted mucosal immune priming and increased microbial translocation. This process introduces intestinal bacteria and MAMPs into the systemic circulation, contributing to cirrhosis-related inflammation, particularly in advanced disease stages. Studies in CLD have reported reductions in commensal bacterial genera such as *Ruminococcus, Faecalibacterium, Bacteroidetes,* and *Coprococcus* (Da Silva et al., 2018; Dubinkina et al., 2017), accompanied by increases in pathobiont taxa like *Enterococcaceae*, *Staphylococcaceae*, and *Enterobacteriaceae* (Bajaj et al., 2014). Additionally, patients with CLD show altered bile acid homeostasis, characterised by alterations in bile acid quantities and composition, which correlate with microbiota shifts and reduced conversion of primary to secondary bile acids (Kakiyama et al., 2013; Mouzaki et al., 2016), with more primary bile acids being found in faeces (Kakiyama et al., 2014). These changes, mirrored in serum bile acids, worsen with cirrhosis progression (Horvatits et al., 2017). However, comprehensive studies integrating microbiota alterations to systemic metabolites, inflammatory and gut barrier markers, across CLD stages remain limited. (Bajaj et al., 2022, 2014, 2012).

This study aimed to identify alterations in bacterial genera in the faecal microbiota, as targets for future therapeutic intervention, across a well-phenotyped cohort of patients with ACLF, decompensated cirrhosis, stable cirrhosis, and compared to healthy controls. We aimed to understand some of the potential functional properties in these microbiome changes by comparison with profiling of serum bile acids, urinary and plasma metabolites, and markers of gut inflammation, bacterial translocation and systemic immune activation, as key pathobiological processes within the gut-liver-immune axis. We hypothesised that microbiota perturbations increase with CLD severity. By correlating these microbiota differences with relevant metabolic, inflammatory, immunological and clinical disease markers, we aimed to detect key target bacteria within the gut that are associated with disruption within the gut-liver-immune axis, providing insights into disease pathobiology and informing potential therapeutic targets.

## 2. Results

### 2.1 Participant characteristics

This cross-sectional cohort included patients with cirrhosis who were categorised based on disease stage and severity; stable cirrhosis (SC), decompensated cirrhosis (DC), or acute-on-chronic liver failure (ACLF). A healthy control (HC) group of healthy participants was included for comparison analysis and to establish normal values. Table 1 summarises demographic, clinical and biochemical characteristics of the recruited patients and healthy controls. A total of 153 patients with cirrhosis were included in this study (SC=15, DC=81, ACLF=57) with 40 HCs. Patients with cirrhosis were older than HC with a mean HC age of 36 years (SD: 10 years) and mean liver cohort age of 53 years (SD: 11 years). Predominant aetiologies of cirrhosis included alcohol-related liver disease (ARLD) (SC/DC/ACLF: 60%/70.4%/57.9%) and metabolic-dysfunction associated steatotic liver disease (MASLD) (SC/DC/ACLF: 20%/8.6%/10.5%), respectively. SC, DC and ACLF patients presented with ascites (33.3%/71.6%/96.5%) and HE (6.7%/43.2%/86%) as predominant manifestations of hepatic decompensation, respectively. None of the cirrhosis patients had experienced a variceal haemorrhage in the 7 days prior to recruitment nor had spontaneous bacterial peritonitis at the time of sampling. DC and ACLF patients were more frequently receiving antibiotics acutely 45.7%/91.2%, respectively) compared to SC (20%). There was no difference in use of rifaximin-α (only five participants received rifaximin-α monotherapy as antibiotic treatment), lactulose, non-selective beta-blockers and acid suppression therapies (proton pump inhibitors, H2 antagonists) across the cirrhosis cohorts. Haematological, biochemical and disease severity and prognostic composite scores followed expected patterns. Transplant-free survival at 90 days was 77.8% and 24.6% for the DC and ACLF groups, respectively, with no deaths observed in the SC group.

**Table 1:**
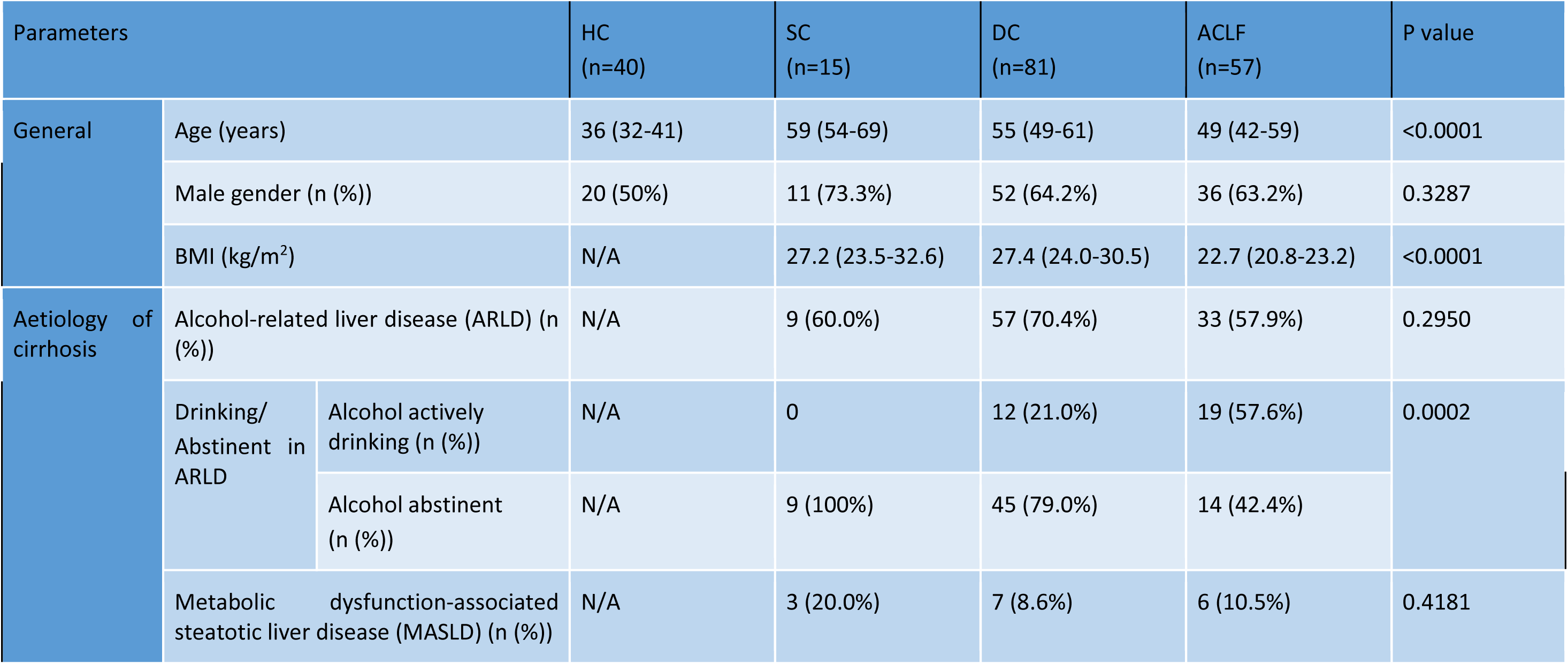
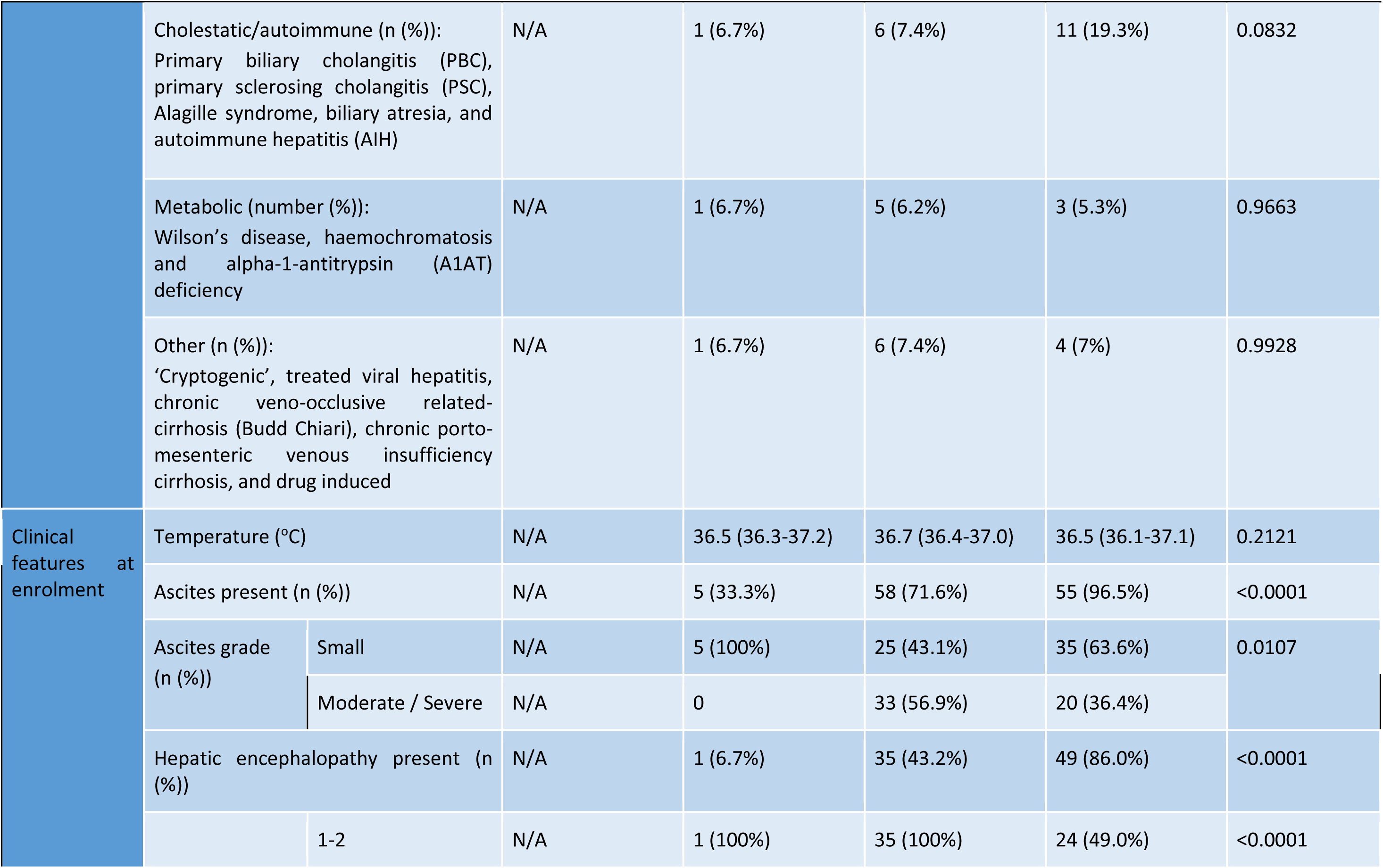
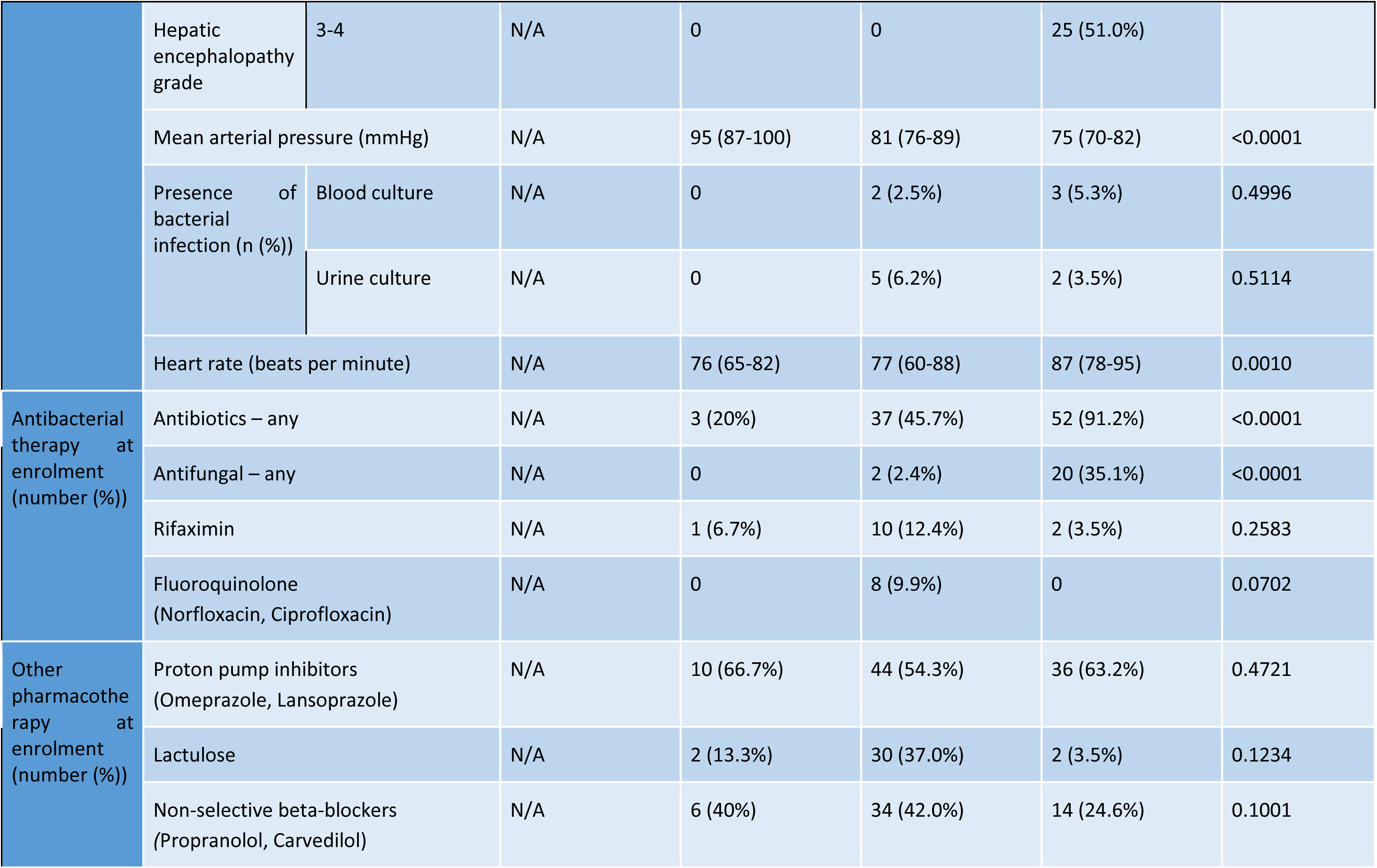
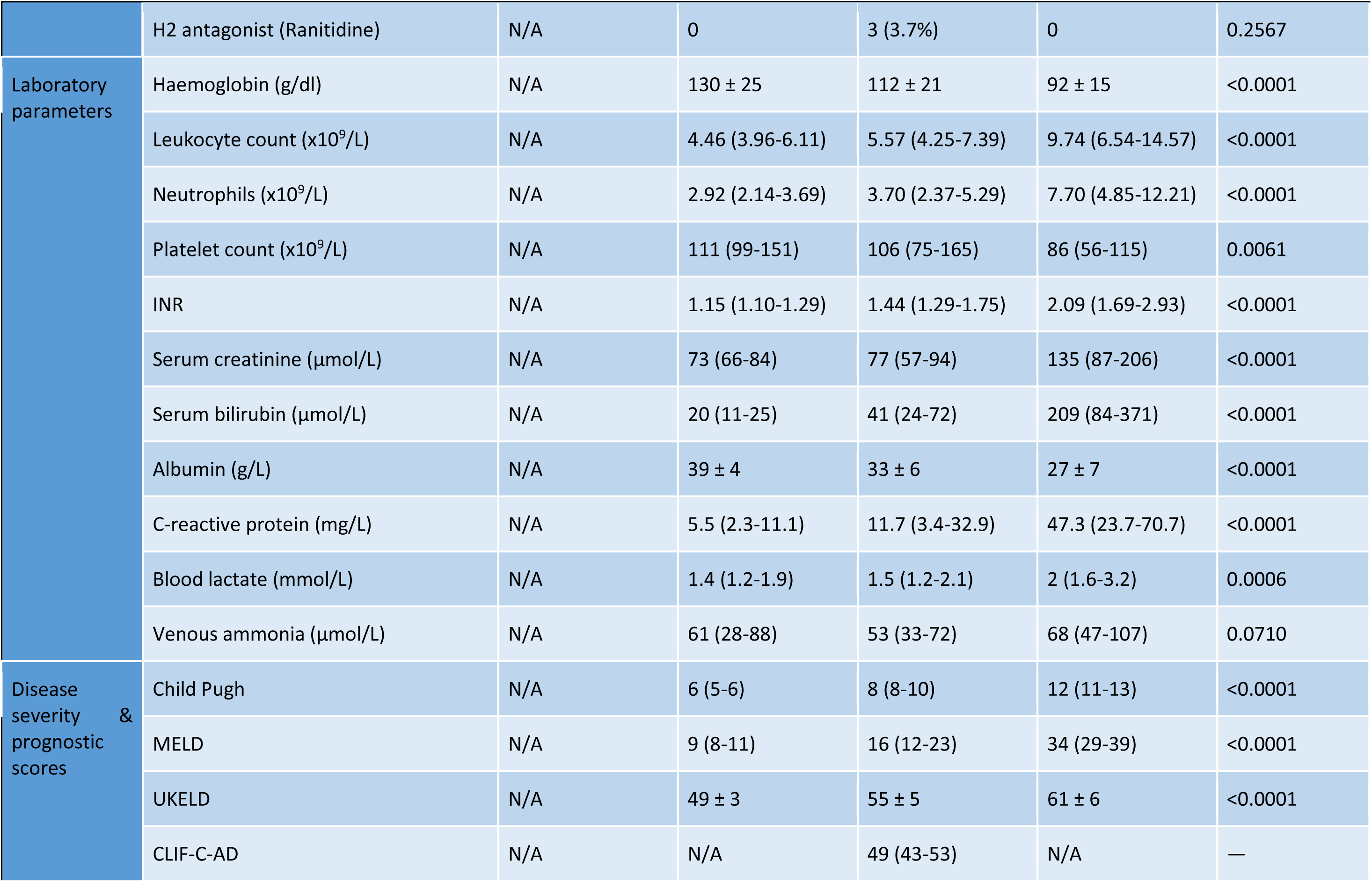
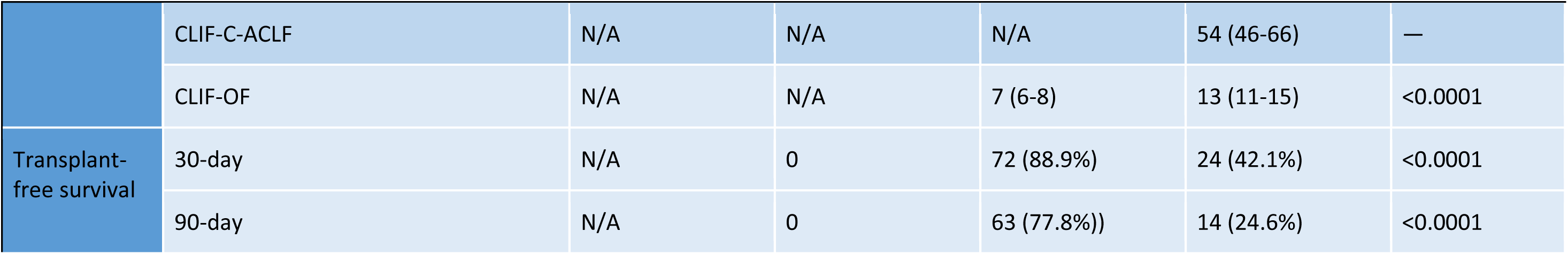
Summary of clinical characteristics of patient and healthy participant study groups (all values given as median [IQR]). Statistics: tatistical analyses were performed using GraphPad Prism, version 10.3.1 (San Diego, USA). Data are presented as medians with interquartile anges [IQRs] or as counts with percentages [%]. Continuous data were analysed with the Mann-Whitney test or Kruskal-Wallis test, as ppropriate. Comparisons between categorical data were conducted using the chi-square or Fisher’s exact test, as appropriate.

| Parameters |  |  | HC<br>(n=40) | SC<br>(n=15) | DC<br>(n=81) | ACLF<br>(n=57) | P value |
| --- | --- | --- | --- | --- | --- | --- | --- |
| General | Age (years) |  | 36 (32-41) | 59 (54-69) | 55 (49-61) | 49 (42-59) | <0.0001 |
|  | Male gender (n (%)) |  | 20 (50%) | 11 (73.3%) | 52 (64.2%) | 36 (63.2%) | 0.3287 |
|  | BMI (kg/m<sup>2</sup>) |  | N/A | 27.2 (23.5-32.6) | 27.4 (24.0-30.5) | 22.7 (20.8-23.2) | <0.0001 |
| Aetiology of cirrhosis | Alcohol-related liver disease (ARLD) (n (%)) |  | N/A | 9 (60.0%) | 57 (70.4%) | 33 (57.9%) | 0.2950 |
|  | Drinking/<br>Abstinent in<br>ARLD | Alcohol actively<br>drinking (n (%)) | N/A | 0 | 12 (21.0%) | 19 (57.6%) | 0.0002 |
|  |  | Alcohol abstinent<br>(n (%)) | N/A | 9 (100%) | 45 (79.0%) | 14 (42.4%) |  |
|  | Metabolic dysfunction-associated<br>steatotic liver disease (MASLD) (n (%)) |  | N/A | 3 (20.0%) | 7 (8.6%) | 6 (10.5%) | 0.4181 |
|  | Cholestatic/autoimmune (n (%)):<br>Primary biliary cholangitis (PBC),<br>primary sclerosing cholangitis (PSC),<br>Alagille syndrome, biliary atresia, and<br>autoimmune hepatitis (AIH) |  | N/A | 1 (6.7%) | 6 (7.4%) | 11 (19.3%) | 0.0832 |
|  | Metabolic (number (%)):<br>Wilson's disease, haemochromatosis<br>and alpha-1-antitrypsin (A1AT)<br>deficiency |  | N/A | 1 (6.7%) | 5 (6.2%) | 3 (5.3%) | 0.9663 |
|  | Other (n (%)):<br>'Cryptogenic', treated viral hepatitis,<br>chronic veno-occlusive related-<br>cirrhosis (Budd Chiari), chronic porto-<br>mesenteric venous insufficiency<br>cirrhosis, and drug induced |  | N/A | 1 (6.7%) | 6 (7.4%) | 4 (7%) | 0.9928 |
| Clinical<br>features at<br>enrolment | Temperature (°C) |  | N/A | 36.5 (36.3-37.2) | 36.7 (36.4-37.0) | 36.5 (36.1-37.1) | 0.2121 |
|  | Ascites present (n (%)) |  | N/A | 5 (33.3%) | 58 (71.6%) | 55 (96.5%) | <0.0001 |
|  | Ascites grade<br>(n (%)) | Small | N/A | 5 (100%) | 25 (43.1%) | 35 (63.6%) | 0.0107 |
|  |  | Moderate / Severe | N/A | 0 | 33 (56.9%) | 20 (36.4%) |  |
|  | Hepatic encephalopathy present (n (%)) |  | N/A | 1 (6.7%) | 35 (43.2%) | 49 (86.0%) | <0.0001 |
|  |  | 1-2 | N/A | 1 (100%) | 35 (100%) | 24 (49.0%) | <0.0001 |
|  | Hepatic encephalopathy grade | 3-4 | N/A | 0 | 0 | 25 (51.0%) |  |
|  | Mean arterial pressure (mmHg) |  | N/A | 95 (87-100) | 81 (76-89) | 75 (70-82) | <0.0001 |
|  | Presence of bacterial infection (n (%)) | Blood culture | N/A | 0 | 2 (2.5%) | 3 (5.3%) | 0.4996 |
|  |  | Urine culture | N/A | 0 | 5 (6.2%) | 2 (3.5%) | 0.5114 |
|  | Heart rate (beats per minute) |  | N/A | 76 (65-82) | 77 (60-88) | 87 (78-95) | 0.0010 |
| Antibacterial therapy at enrolment (number (%)) | Antibiotics – any |  | N/A | 3 (20%) | 37 (45.7%) | 52 (91.2%) | <0.0001 |
|  | Antifungal – any |  | N/A | 0 | 2 (2.4%) | 20 (35.1%) | <0.0001 |
|  | Rifaximin |  | N/A | 1 (6.7%) | 10 (12.4%) | 2 (3.5%) | 0.2583 |
|  | Fluoroquinolone (Norfloxacin, Ciprofloxacin) |  | N/A | 0 | 8 (9.9%) | 0 | 0.0702 |
| Other pharmacotherapy at enrolment (number (%)) | Proton pump inhibitors (Omeprazole, Lansoprazole) |  | N/A | 10 (66.7%) | 44 (54.3%) | 36 (63.2%) | 0.4721 |
|  | Lactulose |  | N/A | 2 (13.3%) | 30 (37.0%) | 2 (3.5%) | 0.1234 |
|  | Non-selective beta-blockers (Propranolol, Carvedilol) |  | N/A | 6 (40%) | 34 (42.0%) | 14 (24.6%) | 0.1001 |

|  |  |  |  |  |  |  |
| --- | --- | --- | --- | --- | --- | --- |
|  | H2 antagonist (Ranitidine) | N/A | 0 | 3 (3.7%) | 0 | 0.2567 |
| Laboratory parameters | Haemoglobin (g/dl) | N/A | 130 ± 25 | 112 ± 21 | 92 ± 15 | <0.0001 |
|  | Leukocyte count (x10<sup>9</sup>/L) | N/A | 4.46 (3.96-6.11) | 5.57 (4.25-7.39) | 9.74 (6.54-14.57) | <0.0001 |
|  | Neutrophils (x10<sup>9</sup>/L) | N/A | 2.92 (2.14-3.69) | 3.70 (2.37-5.29) | 7.70 (4.85-12.21) | <0.0001 |
|  | Platelet count (x10<sup>9</sup>/L) | N/A | 111 (99-151) | 106 (75-165) | 86 (56-115) | 0.0061 |
|  | INR | N/A | 1.15 (1.10-1.29) | 1.44 (1.29-1.75) | 2.09 (1.69-2.93) | <0.0001 |
|  | Serum creatinine (μmol/L) | N/A | 73 (66-84) | 77 (57-94) | 135 (87-206) | <0.0001 |
|  | Serum bilirubin (μmol/L) | N/A | 20 (11-25) | 41 (24-72) | 209 (84-371) | <0.0001 |
|  | Albumin (g/L) | N/A | 39 ± 4 | 33 ± 6 | 27 ± 7 | <0.0001 |
|  | C-reactive protein (mg/L) | N/A | 5.5 (2.3-11.1) | 11.7 (3.4-32.9) | 47.3 (23.7-70.7) | <0.0001 |
|  | Blood lactate (mmol/L) | N/A | 1.4 (1.2-1.9) | 1.5 (1.2-2.1) | 2 (1.6-3.2) | 0.0006 |
|  | Venous ammonia (μmol/L) | N/A | 61 (28-88) | 53 (33-72) | 68 (47-107) | 0.0710 |
| Disease severity & prognostic scores | Child Pugh | N/A | 6 (5-6) | 8 (8-10) | 12 (11-13) | <0.0001 |
|  | MELD | N/A | 9 (8-11) | 16 (12-23) | 34 (29-39) | <0.0001 |
|  | UKELD | N/A | 49 ± 3 | 55 ± 5 | 61 ± 6 | <0.0001 |
|  | CLIF-C-AD | N/A | N/A | 49 (43-53) | N/A | — |
|  | CLIF-C-ACLF | N/A | N/A | N/A | 54 (46-66) | — |
|  | CLIF-OF | N/A | N/A | 7 (6-8) | 13 (11-15) | <0.0001 |
| Transplant-free survival | 30-day | N/A | 0 | 72 (88.9%) | 24 (42.1%) | <0.0001 |
|  | 90-day | N/A | 0 | 63 (77.8%)) | 14 (24.6%) | <0.0001 |

### 2.2. Microbiota diversity, bacterial translocation, faecal calprotectin, serum bile acid, and metabolites profiles are progressively altered with increasing cirrhosis severity

Initial analysis focused on the composition of the faecal microbiota, plasma bile acids, urinary metabolites and serum metabolites between cirrhosis severity groups in comparison to HCs. Faecal microbiota diversity, both the number of bacterial genera detected (**Figure 1A)** and the Inverse Simpson diversity index (**Figure 1B**), was lower in patients with more advanced cirrhosis compared to all other groups. Clustering analysis using non-metric multidimensional scaling (NMDS) shows separation in gut microbiota composition between ACLF and HC, with DC clustered between HC and ACLF, while SC clustered closely to HC (**Figure 1C; Supplementary File 2**). The overall effect of disease severity was tested using a PERMANOVA test (R2 = 13.5%; P = 0.0001) and a pairwise comparison showed that the ACLF group clustered separately (P < 0.05) from DC, SC, and HC groups with DC also different to the HC group (**Supplementary File 6**). To assess the associations between clinical metadata variables, NMDS analysis was specifically conducted on faecal sequencing data from patients with cirrhosis excluding HCs. Envfit analysis was used to examine the influence of 69 clinical variables on microbiota composition. Forty-one clinical variables were significantly associated with microbiota composition in patients with cirrhosis, with measures of cirrhosis severity exhibiting the most pronounced impact (**Supplementary Figure 1A; Supplementary File 7**). For the purposes of visualisation the ten clinical variables with the highest R2 values with the strongest association with the microbiota composition were overlaid on the NMDS plot and were all correlated with cirrhosis severity (**Supplementary Figure 1B; Supplementary File 7**).

**Figure 1:**
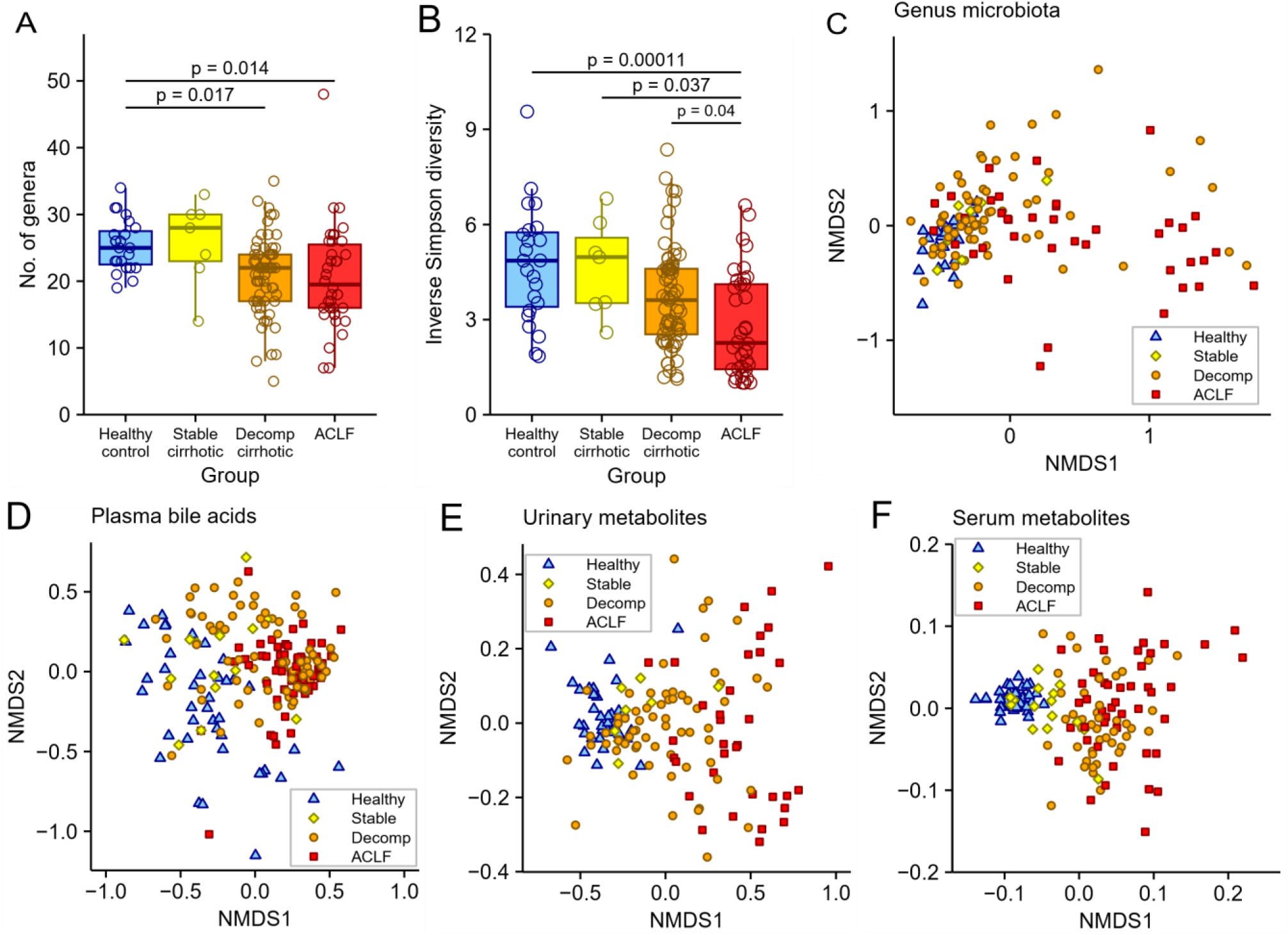
Comparison of faecal microbiota diversity, plasma bile acids, and metabolites profiles in cirrhosis and healthy controls. (A) Number of genera detected in each participant’s faecal sample by cirrhosis severity and HC. (B) Faecal microbiota diversity measured using the Inverse Simpson diversity index by cirrhosis severity vs HC. NMDS ordination shows dissimilarity indices via Bray-Curtis distances between individual participants coloured by cirrhosis severity vs HC (D) plasma bile acids (E) urinary metabolites and (F) serum metabolites. Dot and box plots show median and interquartile ranges of data with circles showing each individual participant.

NMDS plots comparing plasma bile acid composition indicated higher divergence amongst HC samples, and an increasing convergence (clustering) with cirrhosis severity (PERMANOVA: P=0.0001) (**Figure 1D; Supplementary File 3,6**). Both urine (PERMANOVA: P=0.0001) and plasma metabolites (PERMANOVA: P=0.0001) showed significant separation in clustering based on cirrhosis severity with similar NMDS clustering with tightly clustered HC samples and divergent cirrhosis patient samples, particularly for ACLF (**Figure 1E-F; Supplementary File 6**). Multivariate analysis, after obvious exclusions including for dominant glucose or ethanol peaks, for example, showed separation of the four study cohorts (HC, n=36; SC, n=8; DC, n=67; and ACLF, N=36) with pairwise multilevel comparison showing significant differences between all groups except SC and DC (P < 0.05). This was also confirmed visually by the NMDS plots (**Figure E; Supplementary File 5**) and relative metabolite levels per grouping for TMAO, hippurate (**Figure 3F; Supplementary File 6**) and for creatinine, citrate, dimethylamine, formate and mannitol (**Supplementary Figure 3**).

### 2.3. Genus-level microbiota composition at different cirrhosis severities reveals a progressive overabundance of *Enterococcus* with reductions in commensal taxa

To investigate alterations in gut microbiota composition in more detail, bacterial genus profiles within individual participants were examined. Stacked bar plots showing the ten most abundant bacterial genera illustrate an increasing dominance of *Enterococcus* in more advanced cirrhosis i.e. DC and ACLF groups (**Figure 2A**). Differential abundance of bacterial genera is represented in a heat map showing significant associations between faecal bacterial genera comparing the three cirrhosis groups with HC (**Figure 2B; Supplementary file 8**). *Ruminococcus, Roseburia,* and *Lachnospira* showed the strongest negative association with cirrhosis severity with the lowest abundances in ACLF and DC patients compared to HC (**Figure 2C-E**). *Enterococcus* and *Staphylococcus* showed the strongest positive association with cirrhosis severity with highest abundance in ACLF relative to HC (**Figure 2FH**), while *Streptococcus* abundance was higher in DC and SC compared to HC (**Figure 2G)**. *Enterococcus* abundance was markedly higher in the ACLF patient group compared to HC (**Figure 2C)**. Conversely, a proportion of more severely ill patients with cirrhosis have gut microbiota dominated by *Enterococcus*, with significantly elevated levels observed in DC and ACLF. In a quarter of ACLF patients, the relative abundance of *Enterococcus* exceeded 75%.

**Figure 2:**
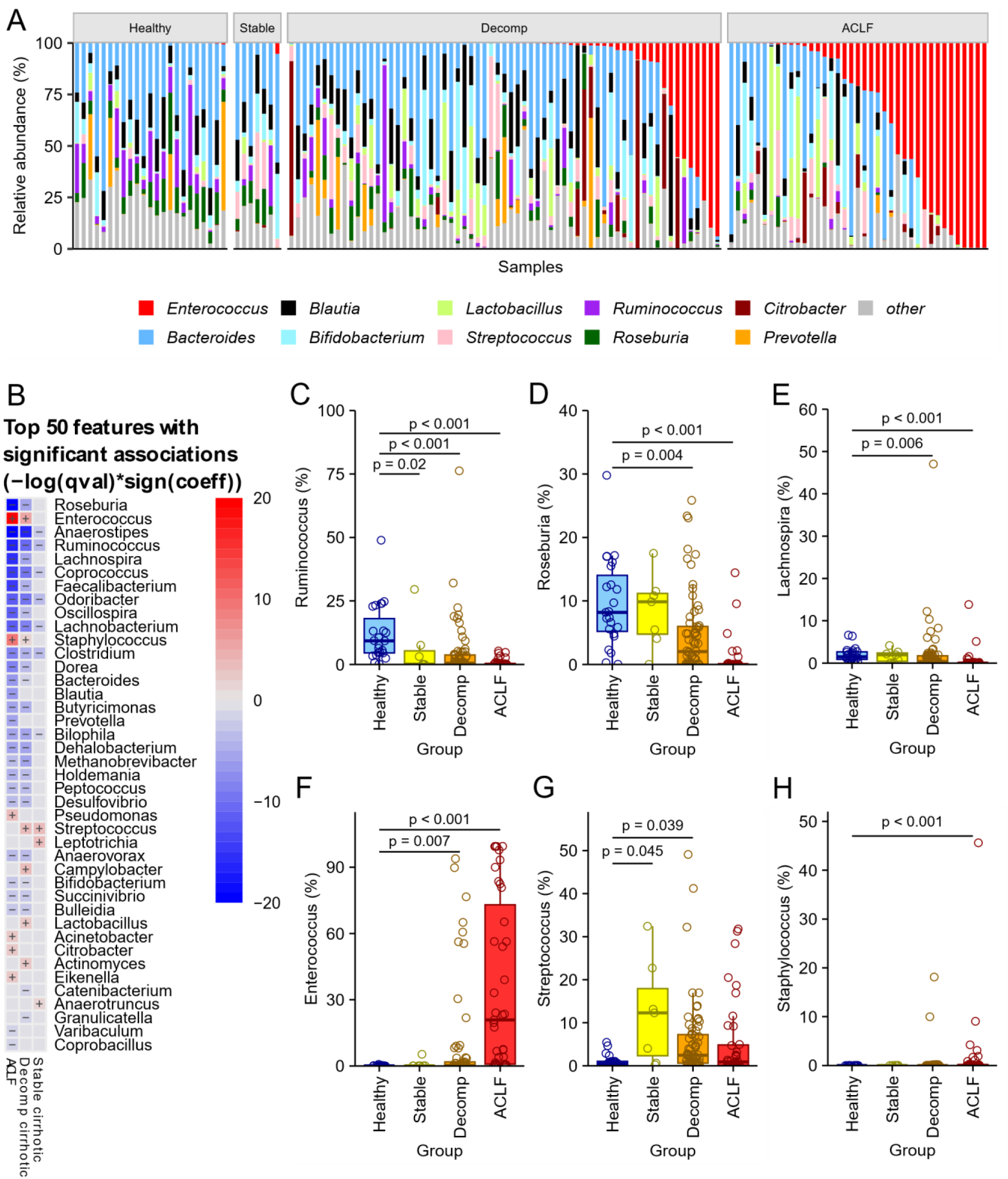
Microbiota genus composition in cirrhosis vs. healthy controls. (A) Stacked bar plots show the ten most abundant bacterial genera in faecal samples stacked by overall abundance, and with participants ordered within cirrhosis severity categories by proportion of *Enterococcus*. (B) The heat map displays MaAslin2 analysis of significant associations between faecal genus abundance and cirrhosis severity with HC used as a reference. The three genera with the strongest negative effect size for cirrhosis severity determined by MaAslin2 using HC as a reference and shown as box and dot plots were (C) *Ruminococcus*, (D) *Roseburia*, and (E) *Lachnospira*. The three genera with the strongest positive effect size for cirrhosis severity determined by MaAsLin2 using HC as a reference and shown as box and dot plots were (F) *Enterococcus*, (G) *Streptococcus*, and (H) *Staphylococcus*. Dot and box plots show median and interquartile ranges of data with circles showing each individual participant.

### 2.4. Plasma bile acids and urinary metabolites correlate with specific gut microbiota taxa with increasing cirrhosis severity

To determine the impact of cirrhosis severity on alterations in plasma bile acids, urinary metabolites, and serum metabolites, we correlated these, using MaAsLin2, with faecal bacterial genera to investigate their associations with gut microbiota composition and provide putative insights into alterations in microbiome function. The primary bile acids taurochenodeoxycholic acid (TCDCA), glycochenodeoxycholic acid (GCDCA), taurocholic acid (TCA) and glycocholic acid (GCA) were higher in the plasma of patients with cirrhosis compared to HC, while other bile acids were lower than in HC (**Figure 3A; Supplementary File 9**). TCDCA and GCDCA showed the strongest effect size with higher concentrations in all three cirrhosis cohorts relative to HC (**Figure 3B-C; Supplementary Figure 3**). Contrasting plasma bile acid concentrations with the percentage of bacterial genera showed that *Enterococcus* abundance was positively correlated with primary bile acids including TCDCA, while *Ruminococcus*, *Coprococcus*, and *Roseburia* were negatively correlated with primary bile acid concentration (**Figure 3D; Supplementary Figure 2**).

**Figure 3:**
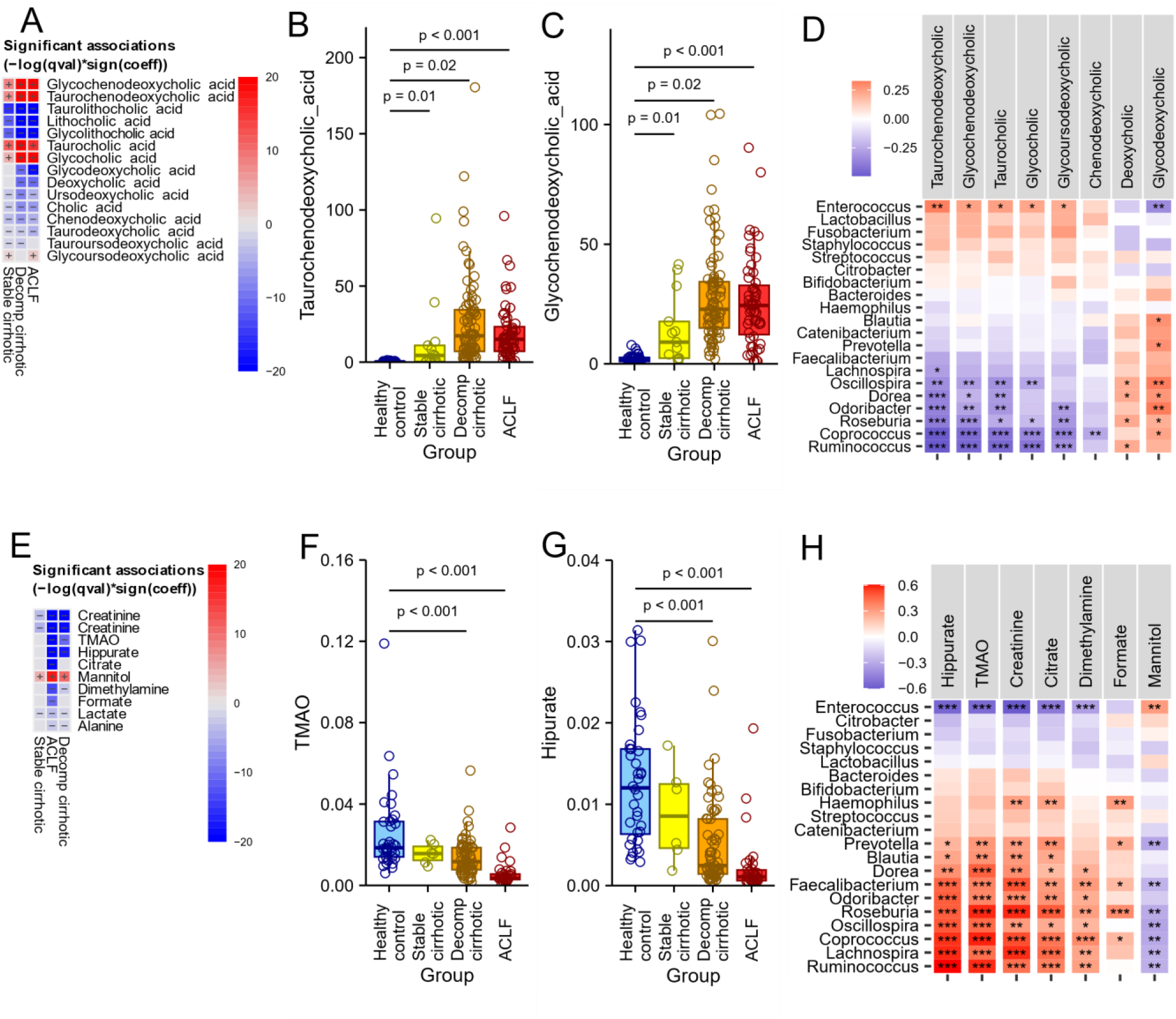
Plasma bile acids, urinary metabolites, and correlations with abundance of faecal bacterial genera. (A) The heatmap displays MaAslin2 analysis results of significant associations between plasma bile acid concentrations and cirrhosis cohorts using HC as a reference. The two plasma bile acids with the strongest positive effect size for cirrhosis severity determined by MaAslin2 using HC as a reference and shown as box and dot plots were (B) Taurochenodeoxycholic acid and (C) Glycochenodeoxycholic acid. (D) Pearson correlations between the normalised relative abundance of the top twenty most abundant faecal bacterial genera (comprising 98% of the microbiota) and normalised plasma bile acid concentrations showing bile acids with significant correlations. (E) The heatmap displays MaAslin2 analysis results of significant associations between urinary metabolite concentrations and cirrhosis severity using HC as a reference. The two urinary metabolites with the strongest negative effect size for cirrhosis severity determined by MaAslin2 using HC as a reference and shown as box and dot plots were (F) Trimethylamine N-oxide (TMAO) and (G) Hippurate. (H) Pearson correlations between the normalised relative abundance of the top twenty most abundant faecal bacterial genera (comprising 98% of the microbiota) and normalised urinary metabolite concentrations showing metabolites with significant (P < 0.05) correlations. Full correlations shown in Supplementary Figure 3 and 4.

Analysis of urinary metabolites showed several metabolites differing by cirrhosis severity relative to HC (**Figure 3E; Supplementary File 10**). Two metabolites, hippurate (**Figure 3F**) and Trimethylamine N-oxide (TMAO) (**Figure 3G**), were identified as products of gut microbial metabolism and both were lower in patients with cirrhosis compared to HC, and lower in ACLF than in SC or DC (**Figure 3F-G**; **Supplementary Figure 2**). Contrasting urinary metabolites with the percentage proportion of bacterial genus showed that *Enterococcus* abundance was negatively correlated with several metabolites including hippurate and TMAO, while *Ruminococcus*, *Coprococcus*, and *Roseburia* were positively correlated with hippurate and TMAO (**Figure 3H; Supplementary Figure 3**).

Similar differences were seen in plasma metabolites with significant differences between cirrhosis cohorts and HC (**Supplementary Figure 4A; Supplementary File 11**) amongst several plasma metabolites (**Supplementary Figure 4B-G**). Similarly, *Enterococcus* abundance correlated with changes in a number of plasma metabolites (**Supplementary Figure 4H**).

These findings show a pattern characterised by elevated concentrations of primary bile acids in plasma and reduced quantities of recognised microbial metabolites in the urine among cirrhosis patients with more advanced disease.

### 2.5. Clinical variables link to overall microbiota profiles, with further associations in advanced cirrhosis of *Enterococcus* with antibiotic usage and resistance profiles

Next, we carried out an exploratory analysis to determine how clinical parameters might be associated with gut microbiota composition, using the envfit function, focusing on each clinical variable and overall microbiota beta-diversity. The ten clinical variables with the strongest association with faecal microbiota composition were plotted overlaying an NMDS plot (**Supplementary Figure 1**). Several clinical factors correlated with microbiota composition, with conventional measures of cirrhosis severity showing the strongest association (**Supplementary Figure 1, Supplementary File 7**). Composite clinical scores are used to classify cirrhosis severity and for prognostication. Of those included in this analysis, ACLF grade, MELD score, Child-Pugh grade and score, and UKELD score were all associated with microbiota composition, in keeping with the study phenotyping of the cirrhosis cohorts, i.e. SC, DC, and ACLF.

Antibiotic use, oral food intake, and lactulose use were clinical variables identified as both having an association with microbiota composition, and considered likely to have a direct influence on the gut microbiota **(Supplementary File 2)**. Differential abundance testing was carried out using MaAslin2 for antibiotic use, oral food intake, and lactulose use along with cirrhosis severity was included within the MaAslin2 model to adjust for covariance (**Supplementary File 12**). Antibiotic use, oral food intake, lactulose use, all showed significant effects on differential abundance of genus within the patient cohort when adjusted for covariance. Comparing the genus with the large difference in abundance in cirrhosis patients receiving or not receiving enteral intake, *Roseburia* abundance was lower in patients not receiving nutrition *via* enteral routes (normal oral intake and by enteral tube feeding), while *Enterococcus* abundance was higher (**Figure 4A**). Similarly, *Roseburia* abundance was lower in patients receiving antibiotics, while *Enterococcus* abundance was higher (**Figure 4B**). Results were unchanged when excluding the 5 patients only receiving rifaximin as antibiotic treatment. Lactulose use was associated with a higher relative abundance of *Bifidobacterium* within the patient cohort between those receiving lactulose and those not (Figure 4E). *Enterococcus* remained significantly higher abundance in DC and ACLF severity groups compared to HC when adjusted for antibiotic use, oral food intake indicating an independent effect of all three factors on *Enterococcus* abundance (**Supplementary Table 2**).

**Figure 4:**
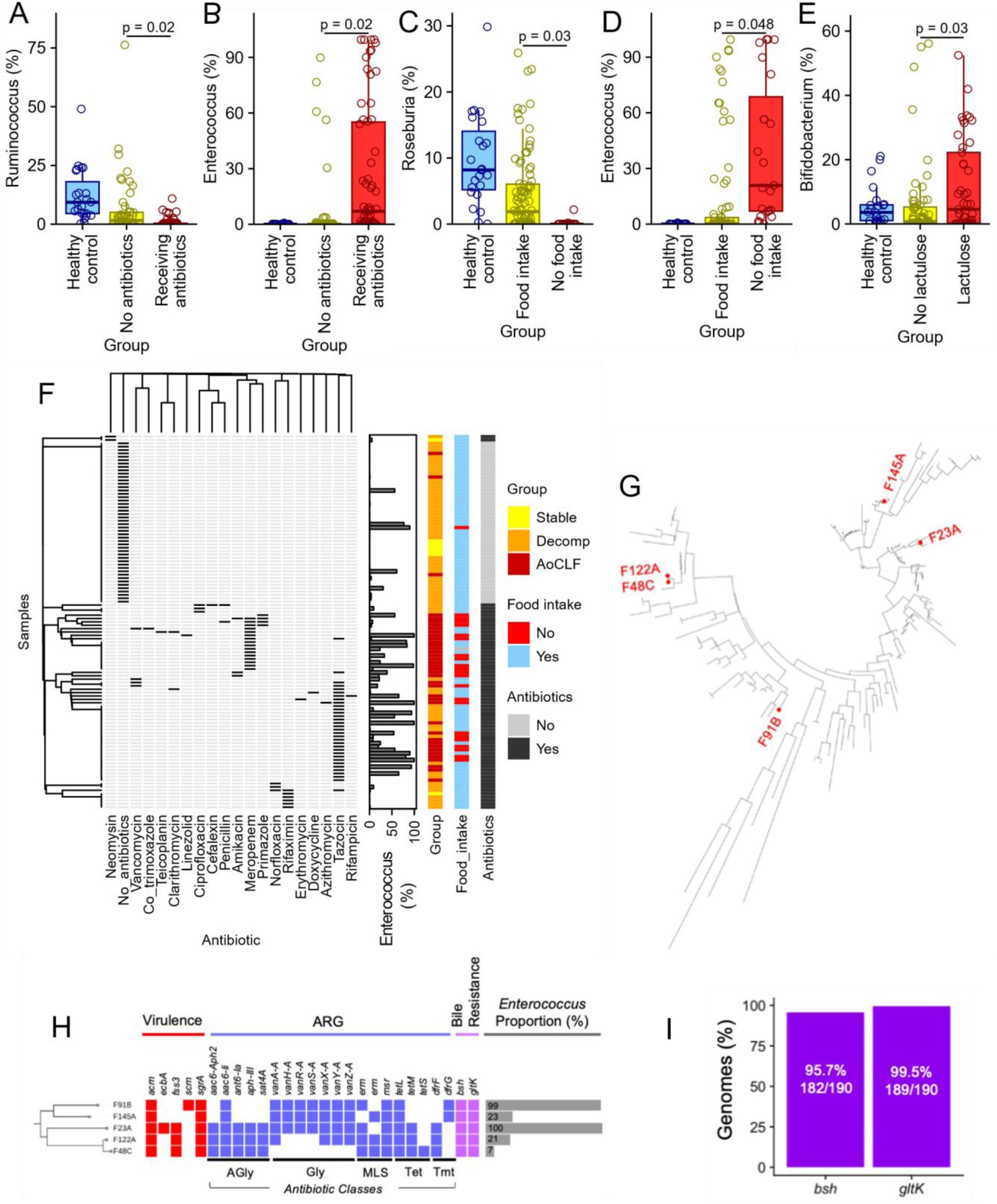
Oral food intake, receiving antibiotics, and *Enterococcus faecium* genomic data. The differentially abundant genera with the strongest negative and positive effect sizes for antibiotics with patients not receiving antibiotics as the reference were (A) *Ruminococcus* and (B) *Enterococcus*. The differentially abundant genera with the strongest negative and positive effect sizes for nutritional intake with patients not receiving enteral as the reference were (C) *Roseburia* and (D) *Enterococcus*. The differentially abundant genera with the strongest positive effect size for lactulose use with patients not receiving lactulose as the reference was (E) *Bifidobacterium*. (F) Clustered individual antibiotics received by patients with cirrhosis cohort, nutritional status, and relative abundance of *Enterococcus* in each sample. (G) A midpoint rooted maximum-likelihood tree consisted of 190 *Enterococcus faecium* genomes, including 5 novel *E. faecium* isolates obtained from cirrhosis patients, as highlighted in the tree. (H) Novel *E. faecium* genomes (n=5) from cirrhosis patients aligned with virulence-related gene profiles, virulence (using VFDB), ARG (Antibiotic Resistance Genes; using ARG-ANNOT) and bile resistance genes (*bsh* and *gltK*), accordingly. Proportion of *Enterococcus* (genus level; based on 16S rRNA sequence data) in the corresponding faecal samples was also indicated in the horizontal bar chart. (I) Proportion of detectable bile resistance genes (%) *bsh* and *gltK* in 190 *E. faecium* genomes, suggesting that these two genes belong to the core genome of *E. faecium*.

The cirrhosis group as a whole received a total of 19 different types of antibiotics (**Figure 4F; Supplementary Table 2.** The most frequently used antibiotics were piperacillin/tazobactam (Tazocin) (33 patients) and 16 Meropenem (16 patients) totalling 37% of patients while 48 (36%) received no antibiotics. The remaining 17 antibiotics were given to 6 or fewer patients with 8 antibiotics only given to one patient. To visualise the complexities of this antibiotic administration, data was visualised as a heatmap with patients clustered by antibiotic use and antibiotics clustered by co-occurrence (**Figure 4C**). Associations of gut bacterial genus differential abundance with all individual antibiotic administration was tested in a MaAslin2 model with only piperacillin/tazobactam (Tazocin) and meropenem individually associated with abundance of *Enterococcus* (**Supplementary Table 8 Figure 4C**). Five out of 48 patients not currently receiving antibiotics also showed higher *Enterococcus* relative abundance (**Figure 4C**). This cirrhosis cohort included a range of aetiologies, however, aetiology did not influence gut microbiota composition. Rather, cirrhosis severity, antibiotic exposure, and nutritional intake were the primary factors that co-associated with a dominant *Enterococcus* gut microbiota signature in patients with the most severe disease i.e. DC and ACLF (**Supplementary figure 7A,B**).

To probe *Enterococcus* traits in more detail, five isolates obtained from five different patient faecal samples (one sample from the DC cohort and four from the ACLF cohort) underwent whole genome sequencing and were identified as *Enterococcus faecium*. Comparing the similarities of these 5 novel isolates with an additional publicly available 185 *E. faecium* genomes (retrieved from the NCBI GenBank database) using midpoint rooted maximum-likelihood tree showed a diversity of *E. faecium* genomes with no single clonal group colonising patients with cirrhosis indicating that the *E. faecium* isolates did not originate from the same source (**Figure 4D**). The five *E. faecium* cirrhosis isolate genomes were aligned against virulence-related gene profiles, antibiotic resistance genes, and bile resistance genes (**Figure 4E**). These isolates contained between 12 and 17 of the 20 antibiotic resistance genes analysed (**Figure 4E**); 5 genes encoding aminoglycoside resistance, 7 *van* genes encoding vancomycin resistance, 3 encoding macrolides resistance, 2 encoding tetracycline resistance, and 2 trimethoprim resistance. The two bile acid resistance genes *bsh* and *gltK* were present in all patient isolates, and in the genomes of 95% of all 190 isolates from public databases (**Figure 4F**).

Taken together, these data indicate that clinical factors such as reduced enteral nutritional intake and exposure to the broad-spectrum beta-lactam antibiotics piperacillin/tazobactam (Tazocin) and meropenem were strongly linked to higher levels of *Enterococcus* and decreased abundance of commensal genera in patients with cirrhosis. Broad-spectrum beta-lactam antibiotics are commonly administered to DC and ACLF patients as part of their treatment regimen. *Enterococcus* isolates obtained and sequenced from these patients were identified as *E. faecium* and were found to encode various antibiotic and bile acid resistance genes, indicating potential bacterial survival mechanisms and pathogenicity.

### 2.6. Faecal microbiota and markers of bacterial translocation, intestinal and systemic inflammation, and monocyte exhaustion

To explore links between gut barrier dysfunction and translocation of intestinal bacteria and/or their products from the gut into the systemic circulation, we determined the number of copies of 16S rDNA in whole blood from cirrhosis patients and HC. The number of copies of 16S rDNA in the blood was lowest in HC in keeping with a degree of ‘physiological translocation’ and displayed a stepwise increase with severity of cirrhosis (**Figure 5A**). To identify associations with the faecal microbiota, we correlated copies of 16S rDNA in the blood against percentages of bacterial genera (**Figure 5B**), with *Enterococcus* most strongly positively correlated (**Figure 5C**), and *Lachnospira* most negatively correlated (**Figure 5C**). Faecal calprotectin as a conventional marker of non-specific gut mucosal inflammation also showed a stepwise increase with cirrhosis severity compared to HC (**Figure 5D**). Faecal calprotectin was positively correlated with relative abundance of *Enterococcus* **(Figure 5E**) and negatively correlated with *Lachnospira* (**Figure 5E**).

**Figure 5.**
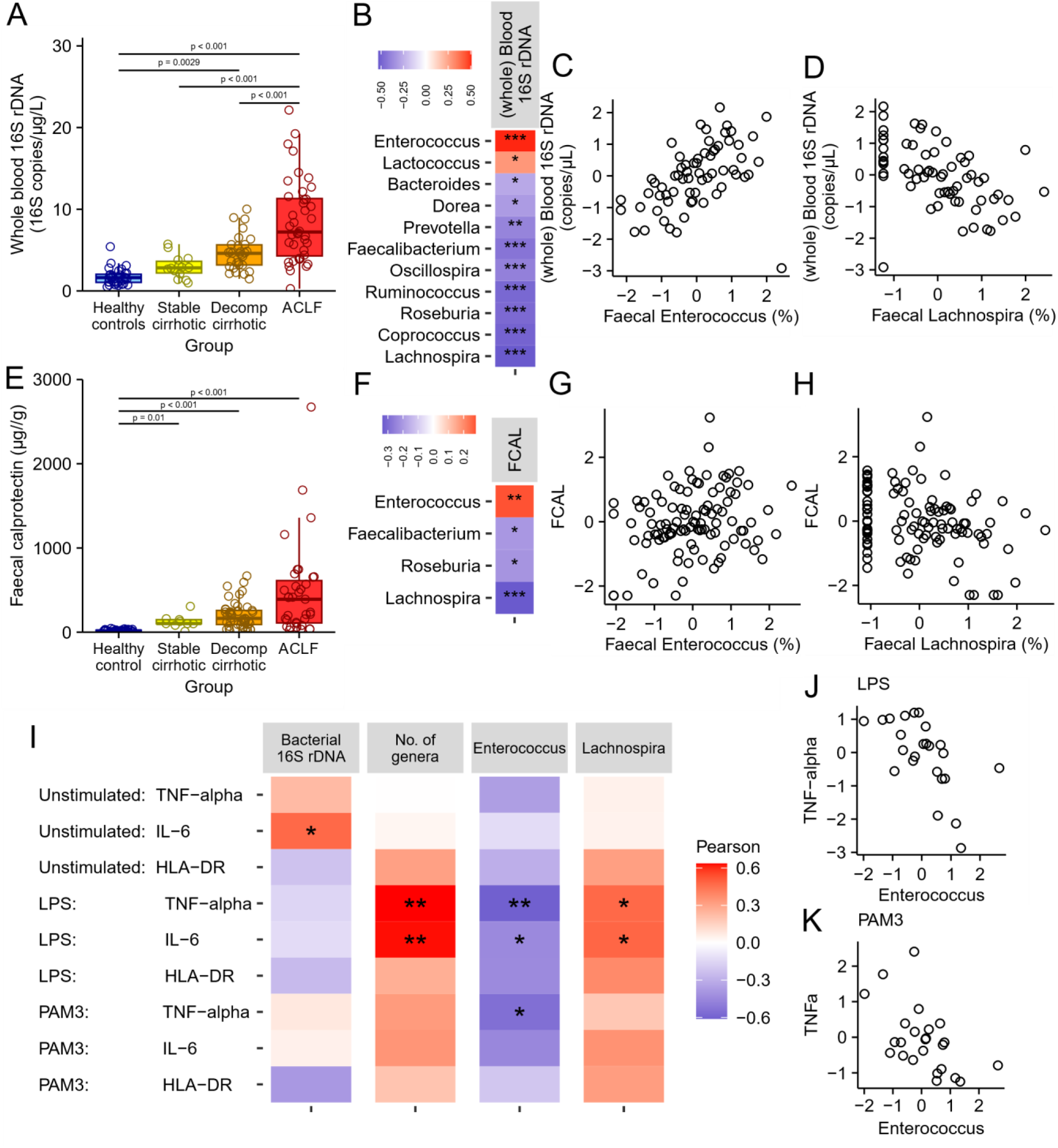
Markers of intestinal permeability and intestinal inflammation and correlations of stimulated monocyte activation with (whole) blood 16S rDNA and faecal microbiota. (A) Copies of 16S rDNA in participant blood by cirrhosis severity vs HC. (B) Significant correlations between copies of 16S rDNA in participant blood and percentage of genera in the faecal microbiota. (C) Scatter plot of copies of 16S rDNA in blood against faecal *Enterococcus* for each participant. (D) Scatter plot of copies of 16S rDNA in blood against faecal *Lachnospira* for each participant. (E) Faecal calprotectin concentration by cirrhosis severity vs HC. (F) Significant correlations between faecal calprotectin concentration and percentage of genera in the faecal microbiota. (G) Scatter plot of faecal calprotectin concentration against faecal *Enterococcus* percentage for each participant. (H) Scatter plot of faecal calprotectin concentration against faecal *Lachnospira* percentage for each participant. (I) Percentage of monocytes showing TNF-α and IL-6 production and HLA-DR expression in response to antigen stimuli (LPS/TLR-4; Pam3CSK4/TLR-2, CpG/TLR-9) correlated separately with whole blood 16S rDNA, number of faecal genera, faecal *Enterococcus* percentage and faecal *Lachnospira* percentage. Stars (*) indicate statistical significance (P < 0.05) after adjustment for multiple testing. (J) Scatter plot of percentage of TNF-α positive LPS stimulated monocytes against faecal *Enterococcus* percentage for each participant. (K) Scatter plot of percentage of TNF-α positive PAM3 stimulated monocytes against faecal *Enterococcus* percentage for each participant.

The innate immune function of CD14+ monocytes isolated from peripheral blood obtained from cirrhosis patients and HCs was assessed by stimulation with TLR 2/4/9 agonists *in vitro*, with unstimulated controls, and the percentage of Tumour necrosis factor-alpha (TNF-α), Interleukin-6 (IL-6), and monocyte human leukocyte antigen-DR (HLA-DR) positive monocytes identified by flow cytometry. Monocytes from DC and ACLF patients showed attenuated proinflammatory responses (TNF-α and IL-6) to Pam3, LPS and CpG challenge, indicating reduced innate responses to pathogen-induced TLR signalling pathways. (**Supplementary Figure 1**). To identify cirrhosis-associated alterations in monocyte function and associations with factors linked to the microbiota, the percentage of TNF-α or IL-6 positive monocytes were correlated with copies of 16S rDNA in whole blood, faecal calprotectin, and the twenty faecal genera with the highest overall abundance (**Figure 5I; Supplementary Figure 6**). The number of copies of bacterial DNA in whole blood positively correlated with the percentage of IL-6 positive unstimulated monocytes, while the number of genera positively correlated with the percentage of TNF-α and IL-6 positive LPS-stimulated monocytes (**Figure 5I**). *Lachnospira* showed positive associations and *Enterococcus* negative associations with the percentage of TNF-α and IL-6 positive LPS stimulated monocytes (**Figure 5J**) and *Enterococcus* with TNF-α positive Pam3 stimulated monocytes (**Figure 5IK**).

These findings reveal contrasting associations between *Enterococcus* and other commensal bacterial genera, such as *Lachnospira*, on markers of intestinal permeability, intestinal and systemic inflammation, and monocyte innate immune function.

## 3. Discussion

We found higher relative abundance of *Enterococcus* in the gut microbiota of patients with cirrhosis, particularly in those with advanced disease and receiving broad spectrum antibiotics, accompanied by a lower abundance of obligate anaerobic commensal genera. This shift is consistent with patterns seen in other conditions with compromised intestinal barrier function, such as preterm infants (Hufnagel et al., 2007) and Graft-versus-Host Disease (Garrett, 2020), as well as other studies in ACLF (Lee et al., 2024a, 2024b; Solé et al., 2021). These findings suggest that *Enterococcus*, an opportunistic pathogen, may exploit disrupted gut microbial communities and diminished colonisation resistance to thrive, especially in advanced disease states with multiple gut microbiota modulating treatments.

Multiple factors may contribute to the selection and proliferation of resident *Enterococcus* strains in the gut of cirrhosis patients. Antibiotic use is one such factor, as many of the patients with high *Enterococcus* levels had received broad-spectrum beta-lactam antibiotics like Piperacillin/tazobactam or meropenem. These antibiotics are commonly administered to hospitalised DC and ACLF patients for treating suspected infections, often before culture results are available, and *Enterococcus* exhibits 100-fold less susceptibility to beta-lactam antibiotics (Zapun et al., 2008), potentially providing a selective advantage. Additionally, the *E. faecium* isolates in this study encoded genes conferring bile acid resistance (Zhang et al., 2013), which may further facilitate persistence in the bile-rich environment characteristic of advanced cirrhosis (Kakiyama et al., 2013). Elevated intestinal luminal bile acids in cirrhosis, particularly primary bile acids, can alter the gut microbial environment (Ridlon et al., 2013), often reducing bile-sensitive commensals like *Roseburia* and providing an ideal niche for *Enterococcus* to overgrow.

This relative overgrowth of *Enterococcus* in the gut microbiota, especially strains with virulence genes like *acm* and *sgrA* (encoded in all strains isolated from this study cohort), which enhance adhesion and translocation, presents significant clinical implications. These strains may be associated with systemic infections that can exacerbate cirrhosis-related complications, such as progression from a stable state to DC and ACLF (Hendrickx et al., 2009; Nallapareddy et al., 2008). Moreover, bacterial sepsis is a significant complication in patients with cirrhosis, an increasing recognition of the threat of antimicrobial resistance (Piano et al., 2019; Wong et al., 2021). Carriage of multidrug resistant strains (as indicated by our genomic analysis) may contribute to greater clinical complication episodes by way of hepatic decompensation, organ failure and mortality rates, due to increased translocation of *Enterococcus* via a dysfunctional and inflamed gut barrier into the bloodstream. Furthermore, as *Enterococcus* appears to dominate more severely ill patients to the exclusion of other potential pathogens, this pattern raises questions about its source. Genomic analysis showed divergent strains of *E. faecium* were present in patients rather than any common clonal lineages, which would be the case if multiple patients had acquired the *E. faecium* from each other or from the same hospital environment. This indicated that the *E. faecium* are more likely to represent resident gut strains already in the patient rather than nosocomial infections. Further investigation involving a larger number of isolates will be valuable to confirm these findings.

As faecal *Enterococcus* abundance increased, we observed a corresponding significant decline in key commensal bacterial genera including *Roseburia*, *Ruminococcus*, *Coprococcus*, and *Faecalibacterium*, with many ACLF patients showing an almost complete relative absence. These genera are essential for SCFA production, such as butyrate (Flint et al., 2012), which supports intestinal barrier integrity. The loss of these beneficial microbes is a common feature in cirrhosis (Acharya and Bajaj, 2019) that is linked to the development of gut barrier failure, pathological microbial translocation and cirrhosis-associated immune dysfunction. Their decline appears to be further exacerbated by common therapeutic factors in the treatment of advanced cirrhosis, such as the use of broad-spectrum antibiotics, which impact antibiotic-sensitive species (Zimmermann and Curtis, 2019), and a reliance on fibre-deficient diets, particularly among patients dependent on enteral or parenteral nutrition (present in 45% of the ACLF group). These diets lack the necessary fermentable dietary fibre to sustain SCFA-producing microbes, worsening microbial disruption and potentially contributing to gut permeability.

This microbiota disruption is mirrored by lower urinary levels of metabolites like hippurate which is a products of beneficial microbial metabolism and fibre-rich diets (Lees et al., 2013) and TMAO, which is a microbial byproduct of metabolising nutrients including choline, betaine, and carnitine found in meat, eggs, and fish (Pallister et al., 2017). Hippurate production, often associated with microbial metabolism of phenolic compounds in fruits and vegetables, was positively correlated with *Ruminococcus*, further linking microbial loss with decreased metabolic activity. Similarly, TMAO - produced via hepatic conversion of trimethylamine (TMA), itself a microbial metabolite from dietary sources like carnitine and lecithin - was also reduced, indicating either a reduction in dietary substrates or a loss of the bacteria producing TMA or potentially a loss of liver function to convert TMA into TMAO (van den Berg et al., 2023). Reduced *Ruminococcus* abundance correlated with decreased urinary TMAO, reflecting not only microbial shifts but also dietary limitations and potential hepatic enzymatic changes (FMO1 and FMO3) seen in advanced cirrhosis (Organ et al., 2020) (Gatarek and Kaluzna-Czaplinska, 2021; Williams et al., 2010). These data suggest that microbial and dietary disturbances may jointly contribute to the metabolic and immune challenges observed in cirrhosis progression.

Further, we noted that patients with cirrhosis exhibited elevated plasma bile acids, especially glycochenodeoxycholic and TCDCA, which correlated with cirrhosis severity. This increase likely stems from impaired hepatic bile acid uptake and reduced conversion to secondary bile acids due to the loss of deconjugating genera like *Lachnospiraceae* and *Ruminococcaceae* genera that were significantly depleted in this cirrhosis cohort (Vital et al., 2019). This impaired conversion could further favour the persistence of *Enterococcus*, as elevated primary bile acids can disrupt the gut microbial ecosystem and inhibit commensal bacteria.

Consistent with microbial translocation in cirrhosis, we observed elevated copies of 16S rDNA in the systemic circulation of patients, correlating with cirrhosis severity and reflecting gut barrier dysfunction (Albillos et al., 2022). This microbial translocation, along with elevated faecal calprotectin concentrations, and associated breakdown of gut barrier function and integrity in cirrhosis (Riva et al., 2020), linking with loss of key butyrate-producing microbes like *Ruminococcus* and *Faecalibacterium* genera in DC and ACLF patients (Fagan et al., 2015) (Bach Knudsen et al., 2018). Reduced faecal short-chain fatty acids have been previously reported (Jin et al., 2019; Wang et al., 2023), and a reduced capacity to ferment dietary fibre in cirrhosis patients compared to healthy controls (Jin et al., 2019). *Enterococcus*, which correlated strongly with 16S rDNA copies in the blood, and gut inflammation markers like faecal calprotectin, indicates its likely role in gut inflammation and immune activation in these patients. In cirrhosis, the progressive loss of intestinal barrier integrity allows MAMPs, and viable bacteria, like *Enterococcus*, to translocate into the bloodstream, triggering systemic inflammation and further impairing immune function (Tilg et al., 2022). Indeed, previous work in mice indicates that *E. faecium* triggers colonic cytokine expression, which appears to be strain dependent (Seishima et al., 2019) and related to elevated faecal cytokine levels and other markers of gut barrier dysfunction in DC patients (Riva et al., 2020). Conversely, *Lachnospira* were markedly reduced in cirrhosis patients (Abdugheni et al., 2022), potentially resulting in the loss of the protective role of SCFAs that dampen mucosal immune activation such as through activation and maturation of T regulatory cells (Y et al., 2013). Coupled with impaired immune responses to pathogen-associated TLR signalling, as observed in the CD14+ monocytes of cirrhotic patients, this suggests a vicious cycle of gut perturbations, immune dysfunction, and bacterial translocation that exacerbates systemic inflammation.

Interestingly, despite the widespread antibiotic use in the DC and ACLF patients, *Bifidobacterium* persisted in the microbiota of patients with cirrhosis. This genus is typically considered sensitive to antibiotic use (Moubareck et al., 2005), so its resilience here may be due to the frequent use of lactulose, a non-absorbable disaccharide laxative often prescribed to prevent or treat hepatic encephalopathy. Lactulose acts as a prebiotic, providing a substrate that *Bifidobacterium* can metabolise, thereby promoting its growth. Recent studies have shown lactulose can elevate *Bifidobacterium* levels in cirrhosis patients, which inhibited the antibiotic-resistant pathobiont vancomycin-resistant *E. faecium* through potential mechanisms including lowering the luminal pH and via acetate production (Odenwald et al., 2023). Our results add to these recent findings that lactulose supplementation could help support *Bifidobacterium* populations in patients with cirrhosis (Odenwald et al., 2023).

From a therapeutic perspective, restoring the gut microbiota toward a healthier balance, or “eubiosis”, could mitigate the cycle of immune dysfunction and prevent cirrhosis-related complications (Tranah et al., 2021). Dietary or microbial interventions, such as prebiotics, probiotics, or microbiome-targeted treatments, could offer a promising approach. While faecal microbiota transplantation is currently being trialled (e.g. ClinicalTrials.gov: NCT02862249, NCT06461208, NCT04932577), it carries risks for patients with advanced cirrhosis, where highly personalised or targeted microbiome interventions may be safer (Cibulková et al., 2021) and use of Cefepime to protect anaerobes in gut vs Pip Taz / meropenem. Probiotics or prebiotics aimed at restoring beneficial taxa lost in cirrhosis, such as *Roseburia*, could help inhibit *Enterococcus* overgrowth, and reinforce intestinal barrier integrity, and reduce inflammation. However, these approaches are not available yet clinically or commercially, and significant investment and development is needed to bring such novel, next-generation microbiome-modulating therapies to benefit this patient population. This highlights the urgency for better antimicrobial stewardship and the development of rapid diagnostics that can better target potential pathogens while reducing the use of broad-spectrum antibiotics and deescalating to avoid prolonged courses of exposure.

These findings raise the possibility that the overgrowth of *Enterococcus* and loss of commensal microbiota do not merely correlate with disease severity but are also associated with conventional therapies such as the often untargeted use of antibiotics. The absence of a dietary fibre may also contribute to the microbiota disturbances observed, underpinning the crucial role of nutrition in this setting. However, the cross-sectional study design and single time-point sampling limit the ability to infer causality. Longitudinal studies, combined with animal models, could further clarify whether *Enterococcus* contributes to pathological processes in cirrhosis or is an opportunistic response to the disrupted gut environment. Additionally, refined antibiotic strategies, possibly reducing the selective pressure for *Enterococcus*, alongside targeted dietary support to foster beneficial microbes, may represent viable options to improve outcomes for these high-risk patients.

## 4. Methods

### Study participants & biological sampling

Patient participants were stratified and phenotyped according to clinically relevant groups based on the severity and time course of their underlying cirrhosis, degree of hepatic decompensation, and presence and extent of hepatic and extra-hepatic organ failure at the time of sampling. These groups were stable cirrhosis (SC; n=15), decompensated cirrhosis (DC; n=55) and acute-on-chronic liver failure (ACLF; n=57), with a separately recruited healthy control cohort (HC; n=40). SC was defined as having Child Pugh A grade(score) of A(5-6). DC was defined by the acute development of 1 or more major complications of cirrhosis, including ascites, HE, variceal haemorrhage, and bacterial infection. ACLF was defined and graded according to the number of organ failures in concordance with criteria reported in the CANONIC study (Arroyo et al., 2015; Moreau et al., 2013). Main exclusion criteria included pregnancy, hepatic or non-hepatic malignancy (HCC within Milan criteria were included), pre-existing immunosuppressive states, replicating HBV/HCV/HIV infection, and known inflammatory bowel disease. Further details are provided in the Supplementary section.

Patients were consecutively recruited at King’s College Hospital after admission to the ward or when reviewed in the hepatology out-patient clinic, through an observational study and a clinical trial. The observational study was granted approval by the NHS Health Research Authority NRES Committee London Westminster [REC reference: 12/LO/1417] and local sponsor Research and Development department (KCH12-126). Ethical approval for the interventional trial was obtained from NHS Health Research Authority NRES Committee South Central-Oxford C (Bristol) [REC reference: 14/SC/0088] and from the Medicines and Healthcare products Regulatory Agency for Clinical Trial Authorisation [EudraCT number: 2013-004708-20; ClinicalTrials.gov NCT02019784]. The studies were conducted in compliance with the principles of the Declaration of Helsinki (1996), principles of Good Clinical Practice, Research Governance Framework and where relevant, the Medicines for Human Use (Clinical Trial) Regulations. Clinical service users and caregivers were fully consulted and involved in the design of both the study and the trial, and were given opportunities to review and feedback on overall set-up, delivery and practical aspects, including input into development of all participant facing documents. Only baseline pre-intervention data from the clinical trial participants are used in this manuscript. Fully informed consent was obtained from all participants. Some participants eligible for this study were unable to provide informed consent due to cognitive impairment arising from HE and permission from a legal representative was sought.

Patient participants, or their legal representative in the case of incapacitation of a potential participant, provided written informed consent within 48 hours of presentation. Patients were managed according to standard evidence-based protocols and guidelines (European Association for the Study of the Liver., 2018). Patient and Public Involvement and Engagement was undertaken with a patient advisory group to determine acceptability of the studies, provided their perspective on study design, informational material, measures to minimise participation burden and agreeing on a dissemination plan of the findings.

Demographic, clinical, and biochemical metadata were collected at the time of biological sampling. Standard clinical composite scores used for risk stratification and prognostication included the Child-Pugh score (Pugh et al., 1973), model for end-stage liver disease (MELD) (Kim et al., 2008), United Kingdom model for end-stage liver disease (UKELD) (Asrani and Kim, 2010), and Chronic Liver Failure Consortium-acute decompensation (CLIF-C AD) (Jalan et al., 2015). Healthy controls aged >18 years (n = 40) were recruited to establish reference values for the various assays performed. Exclusion criteria for healthy controls were body mass index <18 or >27; pregnancy or active breastfeeding, a personal history of thrombotic or liver disease; chronic medical conditions requiring regular primary or secondary care review and/or prescribed pharmacotherapies; or current use of anticoagulants, platelet function inhibitors, or oral contraceptives, and no exposure to antibiotics in the previous 6 months.

### Faecal, urine and blood sample collection and preparation

Plasma samples for bile acid and cytokine profiling were obtained within 24 hours of admission to hospital. Blood was drawn into ethylenediaminetetraacetic acid (EDTA) containing vacuum tubes (BD Vacutainer, Franklin Lakes, NJ, USA). Plasma fractions were collected from these tubes following incubation at room temperature and centrifugation at 2,000 x g for 10 minutes at 4°C. Plasma aliquots were stored immediately at −80°C until further analysis. Whole blood from EDTA tubes was aliquoted into sterile microtubes for and stored immediately at −80°C for bacterial DNA quantification.

Urine samples were obtained and centrifugated at 2,000 x g for 10 minutes at 4°C, with the urinary supernatant stored in aliquots for subsequent analysis. Faecal samples for 16S rRNA sequencing and calprotectin quantification were obtained within 48 hours of admission to hospital using non-treated sterile universal tubes. Faecal samples were kept at 4°C without any preservative and within 6 hours were aliquoted into 1 gram vials for storage at −80°C. Serum and urine samples were prepared for NMR analyses using standardised protocols (Dona et al., 2014).

### Isolation and quality assessment of faecal bacterial DNA

Bacterial DNA was isolated from all faecal samples obtained. This was performed utilising an ISOLATE II Faecal DNA Kit (Bioline, London, UK; BIO-52082). Faecal samples were thawed from −80°C at room temperature for up to 30 minutes. 150mg of faecal material (crude faecal from each faecal sample) was added directly to a Bashing Beads Lysis Tube (Zymo Research, Orange County, California, USA) and rapidly lysed by bead beating in a vortex, without the use of organic denaturants or proteinases. The DNA was then bound, isolated and purified using spin columns. To optimise the DNA binding step, β-mercaptoethanol (Sigma-Aldrich, Saint Louis, Missouri, USA) was added to eliminate deoxyribonucleases released during cell lysis, by reducing the disulfide bonds within the deoxyribonuclease enzymes. This prevents enzymatic digestion of the DNA during its extraction procedure, increasing the overall yield. The eluted DNA, free of contaminants and enzyme inhibitors, is then used for downstream molecular biology applications including genotyping and sequencing. Extracted DNA concentration and purity were measured using a NanoDrop spectrophotometer (Thermo Fisher Scientific, Australia). Samples with DNA concentrations below the dynamic range of the NanoDrop spectrophotometer were analysed using Qubit High-Sensitivity (HS) dsDNA Assay kit (Life Technologies, Australia).

### Faecal 16S rRNA gene amplicon library preparation and sequencing

Amplicons were generated using the method as described previously (Jervis-Bardy et al. 2015). Full details of the library preparation and sequencing protocol are provided in Appendix section 9.3. In brief, barcoded amplicon libraries for the bacterial community analysis on the Illumina MiSeq platform were generated using degenerate primers targeting the V1 and V3 hypervariable regions of the bacterial 16S rRNA gene and Nextera XT index kit (Illumina, Inc., Victoria, Australia).

Amplicons were generated using fusion degenerate primers 27 F (5’-TCGTCGGCAGCGTCAGATGTGTATAAGAGACAGAGRGTTTGATCMTGGCTCAG-3’) and 519R (5’-GTCTCGTGGGCTCGGAGATGTGTATAAGAGACAGGTNTTACNGCGGCKGCTG-3’) with ligated overhang Illumina adapter consensus sequences in italic text. In brief, the initial PCR reactions were performed on a Veriti 96-well Thermal Cycler (Life Technologies, Australia). The PCR reactions were performed in the following programme: initiation enzyme activation at 95°C for 3 min, followed by 25 cycles consisting of denaturation at 95°C for 30 sec, annealing at 55°C for 30 sec and extension at 72°C for 30 sec. After 25 cycles, the reaction was completed with a final extension of 7 min at 72°C.

The Illumina Nextera XT Index kit (Illumina Inc., San Diego. CA, USA) with dual 8-base indices were used to allow for multiplexing. Two unique indices located on either end of the amplicon were chosen based on the Nextera dual-indexing strategy. To incorporate the indices to the 16S amplicons, PCR reactions were performed on a Veriti 96-well Thermal Cycler (Life Technologies, Australia). Cycling conditions consisted of one cycle of 95°C for 3 min, followed by eight cycles of 95°C for 30 sec, 55°C for 30 sec and 72°C for 30 sec, followed by a final extension cycle of 72°C for 5 min.

Prior to library pooling, the barcoded libraries were quantified using the Qubit dsDNA HS Assay Kit (Life Technologies, Carlsbad, CA, USA). Results from this quantification step (amplicon concentration) were used in downstream processing to eliminate contamination. The libraries were sequenced by 2 × 300 bp paired-end sequencing on the MiSeq platform using MiSeq v3 Reagent Kit (Illumina) at the Flinders Genomics Facility, Adelaide, Australia. All sequence data generated have been submitted to the Sequence Read Archive (Archive 2014).

### 16S rRNA amplicon sequencing analysis

The 16S rRNA amplicon sequencing raw reads are deposited in NCBI SRA under project PRJNA1223169. Sequencing reads were analysed using OTU clustering approach via QIIME v1.9.1 as previously described (Kiu et al., 2019). Briefly, raw paired-end reads were firstly merged using PEAR v0.9.6 with -q 30 option (Zhang et al., 2014). Next, quality filtering is performed with split_libraries_fastq.py (QIIME) with -q 29 to retain high-quality reads (Caporaso et al., 2010). Subsequently, chimera removal was performed via chimera identification (identify_chimeric_seqs.py) and chimera sequence removal script (filter_fasta.py). OTU clustering was run via open reference approach (pick_open_reference_otus.py) which does not discard unassigned reads, based on SILVA_132 16S rRNA database optimised for use with QIIME (Quast et al., 2013). BIOM outputs of taxonomic profiles generated from QIIME were then exported using MEGAN v6 (Huson et al., 2016).

The sequencing output of 135 samples included one sample containing only 405 sequences and this was discarded from further analysis. The remaining 134 samples contained a mean of 13,905 sequences per sample (Standard Deviation 6,497) with a minimum of 2,761 and maximum of 38,637 sequences per sample. Rarefaction curves were generated using the rarecurve function in the vegan package (Oksanen, 2017) in R to confirm acceptable levels of sequencing depth. Raw data was normalised using the Deseq2 (Love et al., 2014) package in R using the varianceStabilizingTransformation function.

### *Enterococcus* isolation, genomic DNA extraction, Whole Genome Sequencing (WGS) and analysis

Selected faecal samples (∼20mg) were subject to serial dilutions in sterile PBS before spread-plating on Slanetz and Bartley agar (Thermo Scientific) for selection of Enterococcus colonies (Slanetz and Bartley, 1957). Subsequently, distinct deep-red coloured colonies, presumably Enterococci, on the agar plates were picked and re-streaked once on fresh Slanetz and Bartley agar after incubation at 40°C for 24h. Re-streaked agar plates were then incubated at 40°C for a further 24h to ensure isolate purity prior to overnight culturing in 10ml Brain Heart Infusion media at 40°C to generate sizable bacterial pellets for subsequent genomic DNA extraction. Genomic DNA extraction on the Enterococcus isolates was performed using FastDNA Spin Kit for Soil (MP Biomedicals) as per manufacturer’s instructions as described previously (Gooch et al., 2021).

Sequencing library was prepared via a modified Illumina Nextera Flex low input tagmentation approach as described previously (Baker et al., 2021; Gooch et al., 2021). Genomic DNA samples were sequenced on Illumina NextSeq 500 with read lengths of 150bp (paired-end).

Sequencing reads were quality-filtered and adapter-removed using fastp v0.20.0 with -q 20 option (Chen et al., 2018). Next, short reads were assembled using Unicycler v0.4.9 via primary de novo assembler SPAdes v3.11.1 prior to read polishing via Pilon v1.22. Contigs of length <500 bp were then removed from all draft genome assemblies (Bankevich et al., 2012; Walker et al., 2014; Wick et al., 2017). Taxonomic assignment (Enterococcus faecium) to novel draft genome assemblies were performed using GTDB-Tk v1.5.1 (Chaumeil et al., 2020).

A total of 185 E. faecium complete genome assemblies were retrieved from NCBI Genome database (December 2021) for comparison purposes. Together with 5 novel *E. faecium* genomes (n=200), a core gene alignment was generated using Panaroo v1.2.8 with--merge_paralogs option while SNP variants were next extracted by snp-sites v2.3.3 (Page et al., 2016; Tonkin-Hill et al., 2020). Subsequently, a phylogenetic tree was reconstructed via FastTree v2.1-gtr (-nt) model based on the alignment of core genome SNPs and visualised on iTOL v6 (Letunic and Bork, 2019; Price et al., 2009).

Gene search was performed via ABRicate v1.0.1 with--mincov=90 and--minid=90, using databases ARG-ANNOT v6 (AMR genes) and VFDB (Virulence genes) (Gupta et al., 2014; Liu et al., 2019). Nucleotide sequences of bile-salt resistance genes bsh (Accession: AY260046.1) and gltK (Accession: EFF34375.1) were retrieved from GenBank (Benson et al., 2018).

### Faecal calprotectin quantification

Faecal calprotectin was measured from frozen faecal samples using a commercially available enzyme-linked immunosorbent assay (Bühlmann Laboratories AG, Schönenbuch, Switzerland) that measures calprotectin in a quantitative manner. This was undertaken in a conventional hospital clinical laboratory (ISO 15189 accredited) with regular interval review of standard operating procedures and equipment calibration. Extraction buffer was added to aliquots of 50mg raw faeces at a weight/volume ratio of 1:50 and homogenised by vortexing for 30 minutes. 2mL of the homogenate was then centrifuged in a microcentrifuge for 5 min at 3000 g. Following centrifugation, the calprotectin ELISA was performed using an automated Grifols Triturus® platform. The ELISA plate is coated with a monoclonal capture antibody highly specific to the calprotectin heterodimeric and polymeric complexes. After incubation, washing, a second incubation with a specific detection antibody, and a further washing step, tetramethylbenzidine (blue colour formation) followed by a stop solution (change to yellow colour) are added by the Triturus. The absorption was determined at an optical density of 450 nm. The linear range of the test was 10-600 μg calprotectin/g faeces with concentration-dependent intra- and inter-assay coefficients of between 2-5% and 4-8%, respectively.

### Whole blood 16S ribosomal DNA quantification

The method for quantifying whole blood 16S ribosomal DNA has been previously published in detail (Vergis et al., 2017). Prior to 16S rRNA gene PCR analysis, frozen whole blood samples were thawed to room temperature over a 15-minute period. Mutanolysin (Sigma, St. Louis, Missouri, USA) was added directly to the whole-blood sample; 10μL of 10U/μL enzyme was added for every 200μL of blood being processed, and the sample was incubated for 30 minutes at 37°C.

DNA isolation was performed using a total whole blood volume of 200μL per sample. 19μL of 10mg/ml lysozyme and 1μL glycogen (Thermo Fisher Scientific; R0561), the latter to aid precipitation, was added to the sample and placed in a water bath at 37°C for 30 minutes. A modified protocol from the QIAmp DNA Blood Mini Kit (Qiagen, Inc., Valencia, CA) was then followed: 200μL of AL buffer was added and the sample vortexed. 20μL of Proteinase K (Thermo Fisher Scientific; EO0491) was then added and then incubated at 56°C for 1 hour in a heat block. After incubation, 200μL of 100% ethanol was added and the resulting lysates were loaded onto the QIAamp DNA Mini kit spin columns.

The columns were then centrifuged at 6,000 x g for one minute, and 500μL wash buffer AW1 added, again centrifuged at 6,000 x g for one minute. 500μL wash buffer AW2 was then added, centrifuged at 13,000 x g for three minutes. 50μL of pre-warmed AE elution buffer was added with 5 minutes waiting time, then centrifuged at 13,000 x g for one minute to obtain the DNA extracts. To maximise extraction, the eluate was reapplied to the spin column and centrifuged at 6,000 x g for one minute. PBS and healthy control whole blood PCR were included to quality check for contamination from the process.

Real-time polymerase chain reaction (PCR) was used to assess the quantity of bacterial DNA by comparing to known standards. The primers and TaqMan probes were the well-characterized RW01 and DG74 primers and TaqMan RDR245 universal probe (synthesized at Applied Biosystems, Foster City, CA) and have been described previously (Jordan and Durso, 2005) with a 380bp 16S rRNA gene target. This target is located in the highly conserved region of 16S bacterial rRNA gene in order to detect the majority of bacterial species found in clinical samples.

The forward primer sequence was: 5′-AAC TGG AGG AAG GTG GGG AT3′ and the reverse primer sequence was 5′-AGG AGG TGA TCC AAC CGC A-3′. The TaqMan probe sequence was 5′-(6-FAM)-TA CAA GGC CCG GGA ACG TAT TCA CCG-(TAMRA)-3′. Each well contained 5.5μL sterile water (UV irradiated), 1μL RW01 forward primer (10uM), 1μL DG74 reverse primer (10μM), 0.5ul RDR245 TaqMan probe (10μM), 10μL of TaqMan Gene Expression Master Mix (Invitrogen; 4369016) and 2μL of sample containing the DNA eluate.

A 96 well plate was used and each sample was run in triplicate, and for each plate that was run blank wells were included, a negative control (sterile PBS) and various healthy control samples spiked with a known quantity of E.coli and then serially diluted to generate a standard curve to allow absolute quantification to be calculated. Accordingly, for each PCR run, a standard curve was generated by including a healthy control blood sample which was spiked with known quantities of E.coli (0.4ng, 0.04ng, 0.004ng, 0.0004ng, 0.00004ng) using serial 10-fold dilutions and a negative control, also all run in triplicate, and were put through the same DNA extraction process. Any sample displaying a positive signal at or below the level of the negative control was considered negative. Any triplicate group with readings >1 copy cycle apart was considered unreliable and discarded; otherwise, the mean reading was calculated.

A StepOnePlus PCR instrument (Invitrogen) was used for real-time PCR, with an initial 2 minutes at 50°C and 10-minute denaturation at 95°C, followed by 40 cycles of 15 seconds at 95°C, 30 seconds at 60C (annealing) and 1 minute at 72°C (extension). Fluorescence measurements on the PCR instrument were collected during the extension phase of each cycle, and a cycle threshold (C_T_) value for each sample and control (including a blank well) was calculated by determining the point at which the fluorescence exceeded the threshold limit, which was set automatically. These values were compared with the *E. coli* standards to obtain a relative quantification of bacterial DNA detected in the whole blood and control samples. Standard curves were generated, and concentrations interpolated in Prism, version 7.0 (GraphPad, La Jolla, CA, USA) The results were extrapolated to picograms of bacterial DNA per millilitre of whole blood.

### Plasma cytokine analyses

Plasma cytokines were measured as follows: IL-2, IL-4, IL-6, IL-8, IL-10, IL-12 p70, IL-13, IFN-Ɣ, IL1β and TNF-α using an electro-chemiluminescence platform (Meso Scale Discovery (MSD)). Samples were run in duplicate on U-PLEX Proinflam Combo 1 Human™ plates (Cat# K15049K-1, MSD, Gaithersburg, MD). This was conducted according to the manufacturer’s protocol. The plate was immediately read on a SECTOR(r) Imager (MSD, Gaithersburg, MD).

### Plasma bile acid profiling

Bile acid (BA) profiling was done using an in-house high performance liquid chromatography tandem mass spectrometry (LC-MS/MS) assay modified from published methodology (Tagliacozzi et al., 2003). The method uses an Acentis fusedcore C18 analytical column (15064.6 mm, particle size 2.7 mm, Sigma-Aldrich) on a Jasco LC2000 series HPLC system attached to an API 3200 triple quadrupole mass spectrometer (Applied Biosystems, Cheshire, UK). Mobile phases comprised (A) methanol or (B) deionised H2O, each containing 5 mM ammonium acetate (w/v) and 0.012% formic acid (v/v). Negative ion mass spectra of the fractionated BAs were recorded in multiple reaction-monitoring mode. Data were captured using Analyst Software version 1.4.2 (Applied Biosystems) and quantitation carried out by peak area analysis corrected to internal standards (a2,2,4,4,d4-deoxycholic acid for each of the bile acid chemistries; unconjugated, glycine- and taurine-conjugated). The method was linear between 0.1 and 10 mmol/L for all BAs and their conjugates with coefficients of variation (CV %) of 1.5–6.8% at the lower limit of quantitation (0.1 mmol/L). The inter-assay CV was 3.6–8.0%.

### Plasma ^1^H NMR spectroscopy

All plasma samples were prepared according to the previously published papers by Dona et al. and Trovato et al. (Dona et al., 2014; Trovato et al., 2021)[TOO ADD]. ^1^H NMR spectra were acquired on a Bruker 600 MHz (Avance III) spectrometer, operating at a ^1^H frequency of 600.16 MHz (14.1 Tesla) at 310K using a 5 mm BBI probe and an automated Sample Jet system (Bruker Biospin, Rheinstetten, Germany). Full explanation of processing is given in the Supplementary section.

### Urine ^1^H NMR spectroscopy

Urinary NMR data were acquired at the Centre for Biomolecular Spectroscopy, King’s College London, using a Bruker 600-MHz (AVANCE NEO) NMR spectrometer with a 1H/13C/15N TCI Prodigy (nitrogen-cooled) probe (Bruker, Billerica, USA) as previously described (Bajaj et al., 2019). Pulse-collect NMR data sets were acquired using PURGE water suppression (Le Guennec et al., 2017) and J-resolved NMR sequences. Signal intensities of metabolite regions were obtained using intellibucketing within Wiley KnowItAll® Informatics, Metabolomics Edition v17.0, Wiley Science Solutions, Colorado, USA. NMR spectral regions excluded from the analysis at the outset were 10.0 ppm and 4.5-5.0 ppm (residual water region). Selected intellibucket regions were manually adjusted to incorporate specific metabolite peaks. The binned NMR spectral data outputted from Wiley KnowItAll® Informatics Metabolomics Edition v17.0 were also analysed using MetaboAnalyst v4.0 program (Chong et al., 2018) and exported to the Quadram Institute. Details can be found in the Supplementary section.

### Monocyte intracellular cytokine staining

Monocyte intracellular cytokine staining was undertaken using thawed PBMCs from patients with liver disease and at a density of 0.5 millions cells per ml. Cells were either stimulated with TLR2 (Pam3CSK4 5ug/ml), Invivogen, France), TLR4 (LPS 100 ng/ml), or TLR9 (CpG 10ug/ml) (Invivogen, France) for 4 hours, or left unstimulated. Monocytes were identified using monoclonal antibodies for surface markers: CD14 PE-Cy7, CD16 APC-H7 (BD Biosciences, UK) and HLA-DR PerCP (ThermoFisher Scientific, UK). Intracellular cytokines were measured using protein transport inhibitor containing monensin, permeabilization/fixation buffer and monoclonal antibodies TNF-α APC and IL-6 PE (BD Biosciences, UK). Percentage positivity in CD14+ monocytes compared with isotype controls (APC mouse IgG1; PE rat IgG2a) were assessed by flow cytometry as previously described (Antoniades et al., 2014). The gating strategy that was employed is shown in Supplementary Figure 1.

### Statistical and bioinformatic analyses

For microbiota diversity analysis the number of genera in each sample and the Inverse Simpson diversity was calculated using the specnumber and the diversity functions on genus level taxonomic sequence data in the vegan package (version 2.6-4) in R using raw sequence count data (Oksanen, 2017). Differences in alpha-diversity between cirrhosis severity and HC groups was tested with an ANOVA using the aov function in R. Post-hoc pairwise comparisons were determined using the TukeyHSD function in R.

To visualise clustering of gut bacterial communities between participants non-metric multidimensional scaling (NMDS) based on a Bray-Curtis distance matrix and using the metaMDS function with k = 3 dimensions contained within the vegan package (version 2.6-4) (Oksanen, 2017). The influence of cirrhosis severity group compared to HC on participant microbiota clustering was tested using a PERMANOVA test using the adonis2 function in the vegan package (version 2.6-4) with a total of 10,000 permutations. Pairwise comparison of clustering of the three cirrhosis severity and HC groups was compared using the pairwiseAdonis function in the pairwiseAdonis package (Martinez Arbizu, 2020).

To explore the effect size and significance of associations between clinical metadata variables and beta-diversity of the microbiota within patient samples the envfit function in the vegan package was used to test associations of clinical metadata variables on NMDS ordination calculated using Bray-Curtis dissimilarity after 10,000 permutations. In total, 76 clinical metadata variables were included in the envfit analysis to explore their associations with the gut microbiota composition with three samples excluded during analysis due to missing data (Supplementary File 2). The P-values for combined continuous and factor variables were corrected for multiple comparisons using an FDR adjustment (Benjamini–Hochberg procedure). For visualisation the R2 for the forty significant variables were presented as a bar plot while an arbitrary top ten clinical variables explaining the highest amount of variance (R2) were plotted as an overlay on a NMDS plot of the individual gut microbiota data using the ggplot2 package.

Associations between the relative abundance of individual bacterial genera and individual clinical metadata variables were tested using the MaAsLin2 package (Mallick et al., 2021). For genus abundance analysis MaAsLin2 was run with default parameters including multiple testing correction using Benjamini-Hochberg FDR with a significance threshold on adjusted P-values of 0.25 as recommended by the developers. Exceptions to default parameters included an applied minimum prevalence of 0.01 and abundance cut-off of 0.01. Each metadata variable was tested individually. For genus differential abundance analysis cirrhosis severity was included at a Fixed Effect with the HC group as the reference group. For genus differential abundance analysis using antibiotic use, oral food intake, and lactulose use each variable was tested individually. Positive associations were identified antibiotic use, oral food intake, and lactulose and these three variables where combined and tested together as Fixed Effects in a multivariable analysis for the final analysis.

Differential analysis of bile acid concentrations was performed using MaAsLin2 with multiple testing correction using Benjamini-Hochberg FDR with a significance threshold on adjusted P-values of 0.25 as recommended by the developers. Exceptions to default parameters included no minimum concentration nor abundance cut-off, CLR as the normalisation method and no transformation used. Differential analysis of urinary metabolite and plasma concentrations were analysed using MaAsLin2 run with default parameters including multiple testing correction using Benjamini-Hochberg FDR with a significance threshold on adjusted P-values of 0.25 as recommended by the developers. Exceptions to default parameters included no minimum concentration nor abundance cut-off, and no normalisation nor transformation used. The BestNormalize package (version 1.9.1.9000) (Peterson, 2021; Peterson and Cavanaugh, 2020) in R was used to optimally normalise each metabolite individually before analysis using the bestnorm function. The method used to transform each variable included in (Supplementary Table 3).

For correlation analysis the variables were normalised using the bestnorm function from the bestNormalize package (version 1.9.1.9000). The method used to transform each variable included in (Supplementary Table 4). Pearson correlations were used with statistical significance corrected for multiple testing across genus tested using the False Discovery Rate method.

## Acknowledgements

We are grateful to all the patient participants and healthy volunteers for agreeing to take part in this study, and to the clinical and liver research teams at King’s College Hospital for facilitating recruitment, collecting metadata and biological sample collection. We are grateful to Dr Mark McPhail for methodological guidance and expertise on plasma NMR data generation. We thank the King’s College Hospital Charity and a generous donation from a Liver Service User, and the Foundation for Liver Research for funding. Sequencing analysis was supported in part by the NBI Research Computing through the provision of a High Performance Computing. The authors thank the Centre for Biomolecular Spectroscopy funded by the Wellcome Trust and British Heart Foundation (ref. 202767/Z/16/Z and IG/16/2/32273).

## Funding statement

The observational study was adopted to the National Institute for Health Research (NIHR) Clinical Research Network (CRN) portfolio, supporting participant screening and recruitment by the Liver Research and Anaesthetics, Critical Care, Emergency and Trauma Teams at King’s College Hospital NHS Foundation Trust. VCP was supported by a New Investigator Award from the Intensive Care Foundation (Registered charity number: 2940178) and Clinical Research Network South London Principal Investigator Strategic “GreenShoot” Research Delivery and Development Funding. The RIFSYS clinical trial was funded through an investigator-initiated study grant awarded by Norgine Pharmaceuticals UK Limited to King’s College London. Infrastructure to support the trial was also provided by the Medical Research Council (MRC) Centre for Transplantation, King’s College London, UK – MRC grant no. MR/J006742/1. Laboratory assays were also part funded by a generous donation from a Liver Service User to the King’s College Hospital Institute of Liver Studies and Transplantation Charity Research Fund, King’s College Hospital Charity (Registered charity number: 1165593). MS, BA, MEM, MM, IJC, SC and VCP were supported and bioinformatic analyses were part funded by the Foundation for Liver Research (Registered charity number: 268211/1134579). LJH is supported by Wellcome Trust Investigator Award (220876/Z/20/Z); the Biotechnology and Biological Sciences Research Council (BBSRC), Institute Strategic Programme Gut Microbes and Health (BB/R012490/1), and its constituent projects BBS/E/F/000PR10353 and BBS/E/F/000PR10356, and the BBSRC Institute Strategic Programme Food, Microbiome and Health BB/X011054/1 and its constituent project BBS/E/QU/230001B. MT Acknowledges support from the NIHR-Imperial Biomedical Research Centre and the Medical Research Council Precision Medicine Award (MR/R014019/1).

## Conflicts of interest

LAE has undertaken consultancy for EnteroBiotix.

DLS has undertaken consultancy for Norgine Pharmaceuticals Ltd, EnteroBiotix, MRM Health, Apollo Therapeutics, Genfit and Satellite Biosciences.

RPV reports Honoraria from Novo Nordisk, Amarin UK and AstraZeneca. AS has delivered paid lectures for Bristol Myers Squibb.

VCP has undertaken consultancy for Norgine Pharmaceuticals Ltd, Alfasigma S.p.A., AstraZeneca and Menarini Diagnostics Ltd.

MD, LH, RK, IJC, ALG, GR, SS, SC, KB have no conflicts.

## Authors contributions

- Matthew J Dalby: Data curation, Formal analysis, Investigation, Software, Visualisation, Writing - Original Draft, Writing - Review & Editing
- Raymond Kiu: Data curation, Formal analysis, Writing - Review & Editing
- Marilena Stamouli: Data curation, Formal analysis, Writing - Review & Editing
- Betsy Arefaine: Data curation, Formal analysis
- Lex E X Leong: Data curation, Formal analysis, Investigation, Visualization, Writing - Review & Editing
- Jack Hales: Methodology, Writing - Review & Editing
- Christine Bernsmeier: Data curation, Formal analysis, Visualization, Writing - Review & Editing
- Arjuna Singanayagam: Data curation, Formal analysis, Visualization, Writing - Review & Editing
- Sidsel Støy: Data curation, Formal analysis, Visualization, Writing - Review & Editing
- Maria-Emanuela Maxan: Data curation, Writing - Review & Editing
- Merianne Mohamad: Visualization, Writing - Review & Editing
- Matt Lewis: Investigation, Methodology, Writing - Review & Editing
- Royce P Vincent: Resources, Writing - Review & Editing
- Jane Cox: Data curation, Formal analysis, Investigation, Visualization, Writing - Review & Editing
- Adrien Le Guennec: Data curation, Visualization, Writing - Review & Editing
- Roger Williams: Supervision
- Lindsey A Edwards: Data curation, Formal analysis, Writing - Review & Editing
- Shilpa Chokshi: Supervision, Writing – review & editing
- Mark Thursz: Resources, Supervision, Writing – review & editing
- Naiara Beraza: Investigation, Resources, Writing - Review & Editing
- Charalambos G Antoniades: Supervision
- Debbie L Shawcross: Conceptualisation, Funding acquisition, Resources, Supervision, Writing – review & editing, served as Chief Investigator of RIFSYS Trial
- Geraint B Rogers: Conceptualisation, Data curation, Formal analysis, Investigation, Resources, Supervision, Visualization, Writing - Review & Editing
- Julia A Wendon: Conceptualisation, Supervision, Writing – review & editing
- Kenneth D Bruce: Conceptualisation, Supervision, Writing – review & editing
- Vishal C Patel: Conceptualisation, Funding acquisition, Investigation, Methodology, Project administration, Resources, Supervision, Writing – review & editing, served as Chief Investigator of GLA study
- Lindsay J Hall: Conceptualisation, Funding acquisition, Investigation, Methodology, Project administration, Resources, Supervision, Writing – review & editing

## Data availability

The 16S rRNA amplicon sequencing raw reads are deposited in NCBI SRA under project PRJNA1223169.

## 7. Supplementary data

**Supplementary Figure 1.**
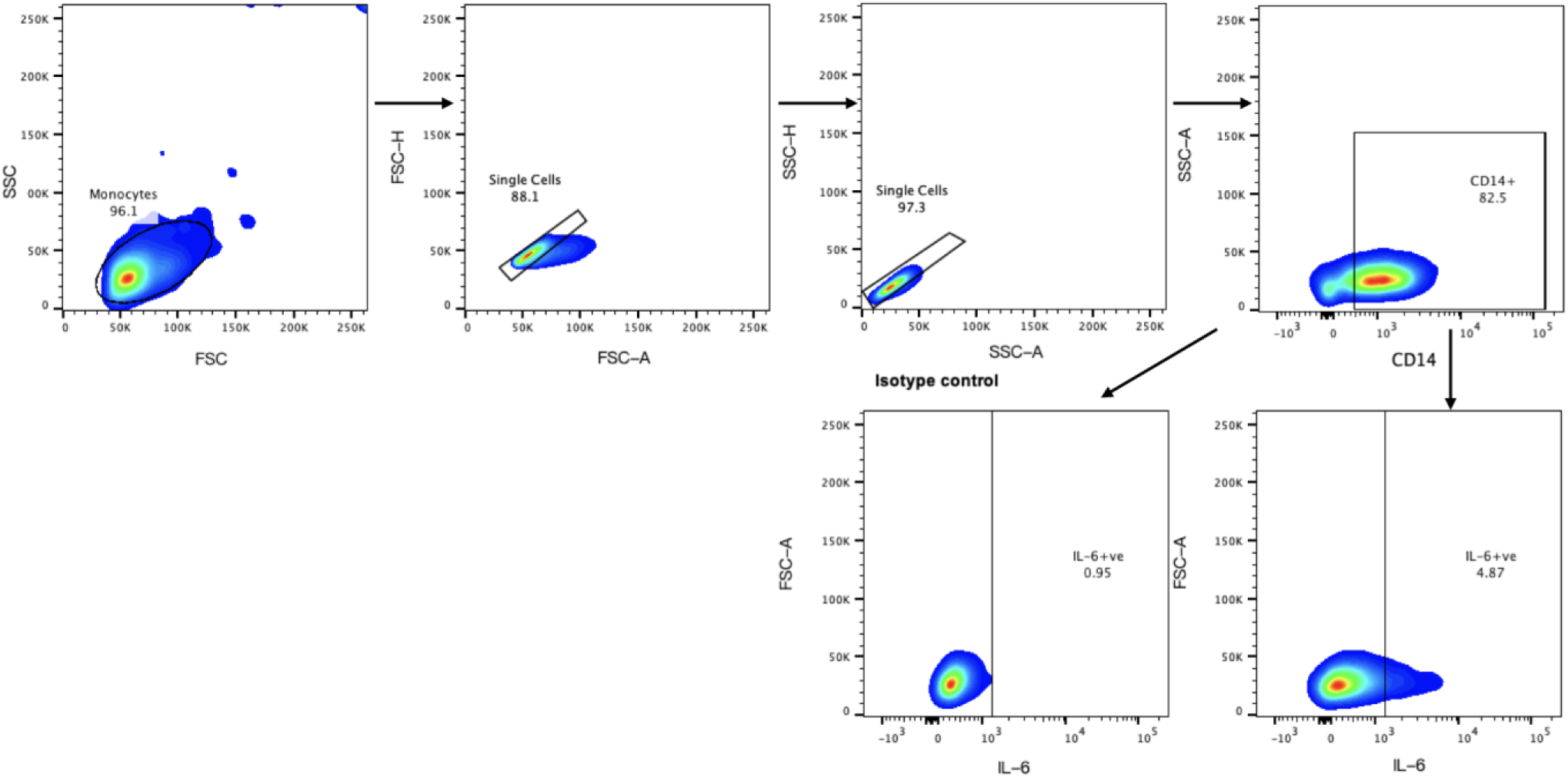
Representative FACS gating strategy used to identify CD14+ monocytes in peripheral blood mononuclear cells (PBMCs) with isotype control used to determine the IL-6-producing monocytes (%) in response to LPS stimulation. Side scatter (SSC), forward scatter (FSC).

**Supplementary Figure 2.**
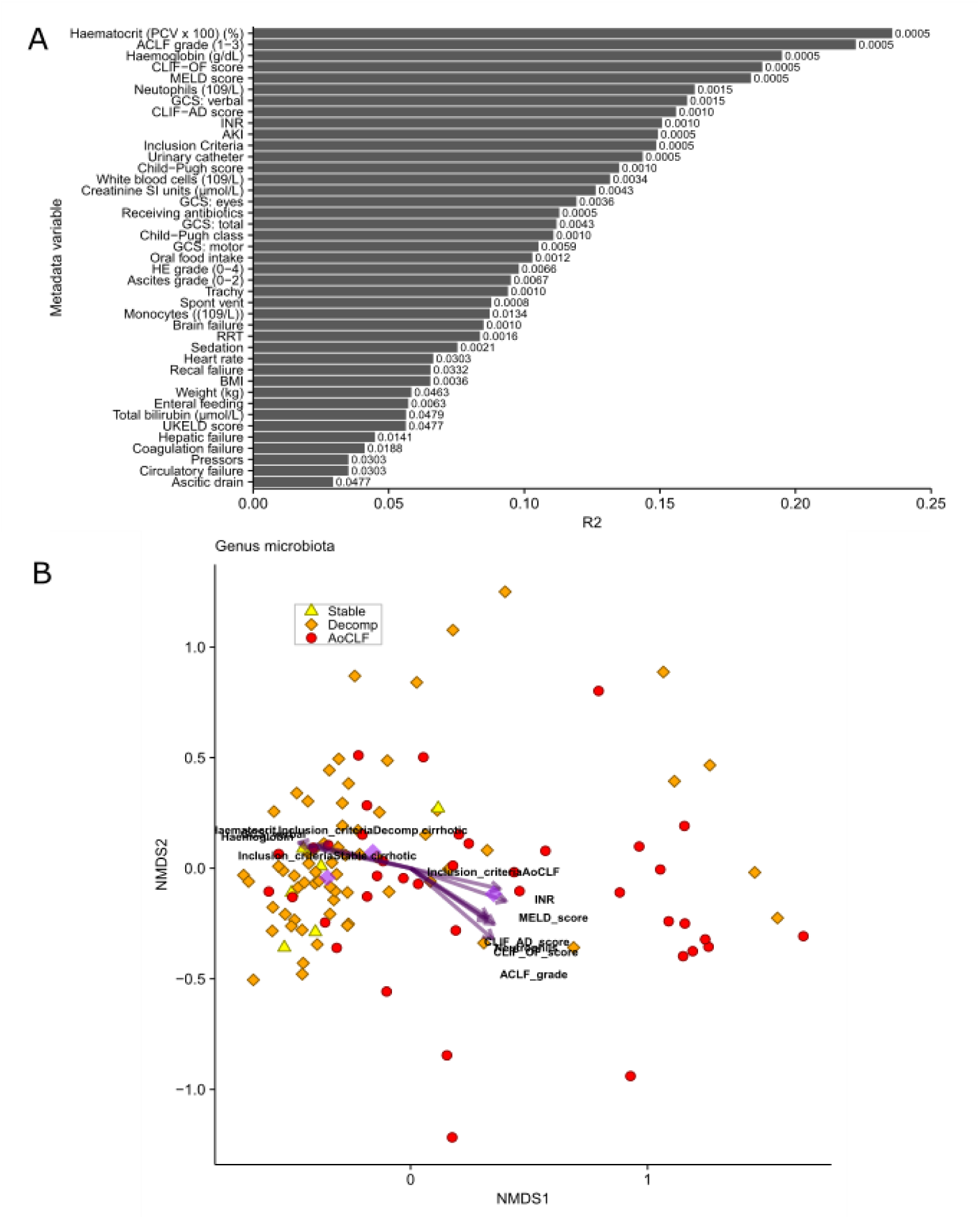
Clinical metadata associated with cirrhosis patient gut microbiota composition. (A) Results of envfit analysis showing clinical metadata variables significantly associated with microbiota composition after correction for multiple testing using False Discovery Rate. Bars show the R2 (squared correlation coefficient) of the correlation and numbers on bars show adjusted P-values. (B) NMDS ordination plot showing gut microbiota composition of individual liver patients with the ten clinical metadata variables with the highest R2 overlayed.

**Supplementary Figure 3.**
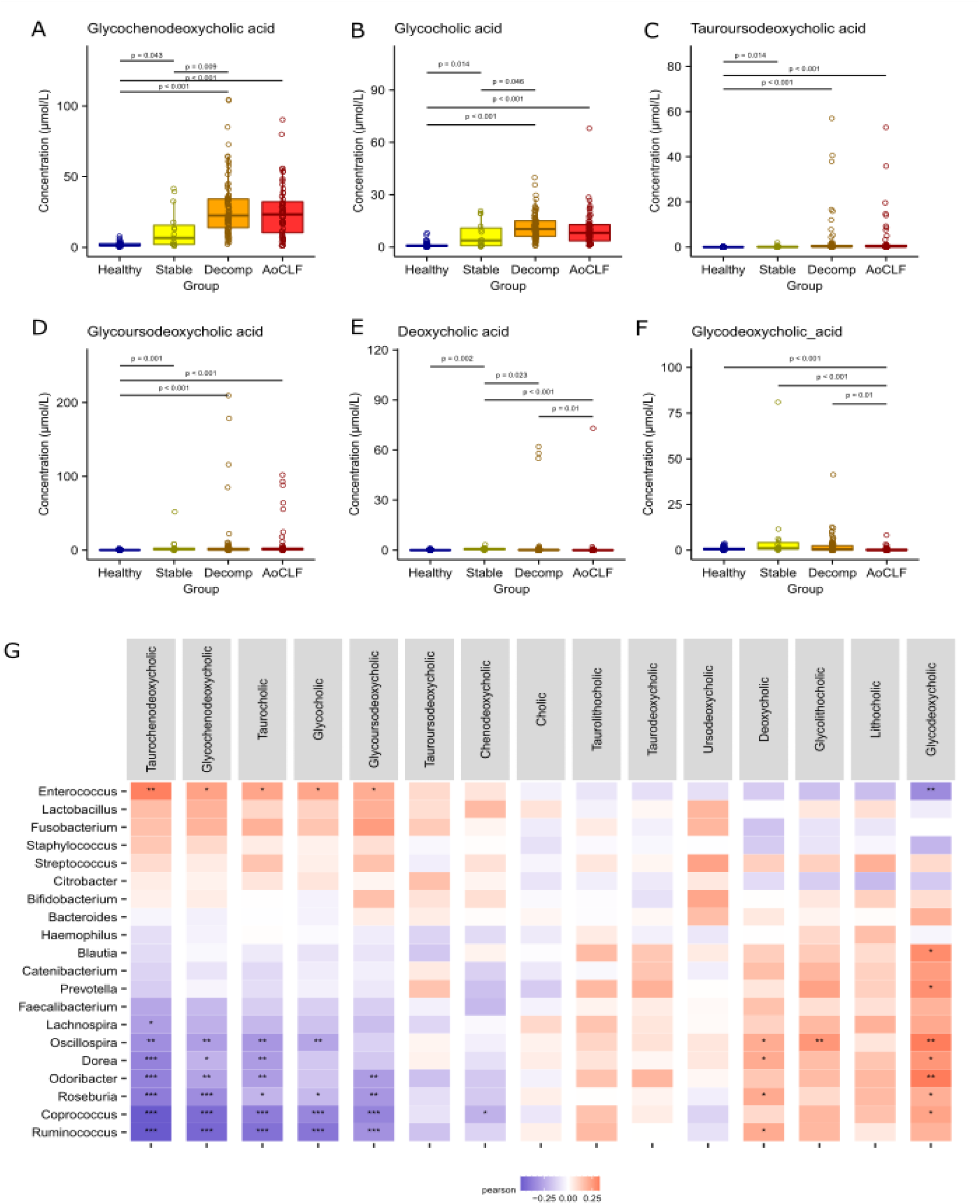
Individual bile acid concentrations by cirrhosis severity and correlations between genus proportion and bile acids. (A) Glycochenodeoxycholic acid, (B) Glycocholic acid, (C) Tauroursodeoxycholic acid, (D) Glycoursodeoxycholic acid, (E) Deoxycholic acid, (F) Glycodeoxycholic acid, (F) Pearson correlations between normalised plasma bile acid concentrations and normalised genus relative abundance using a Pearson correlation. P-value: * = p<0.05, ** = p<0.01, ** = p<0.001. Significance corrected for multiple testing.

**Supplementary Figure 4.**
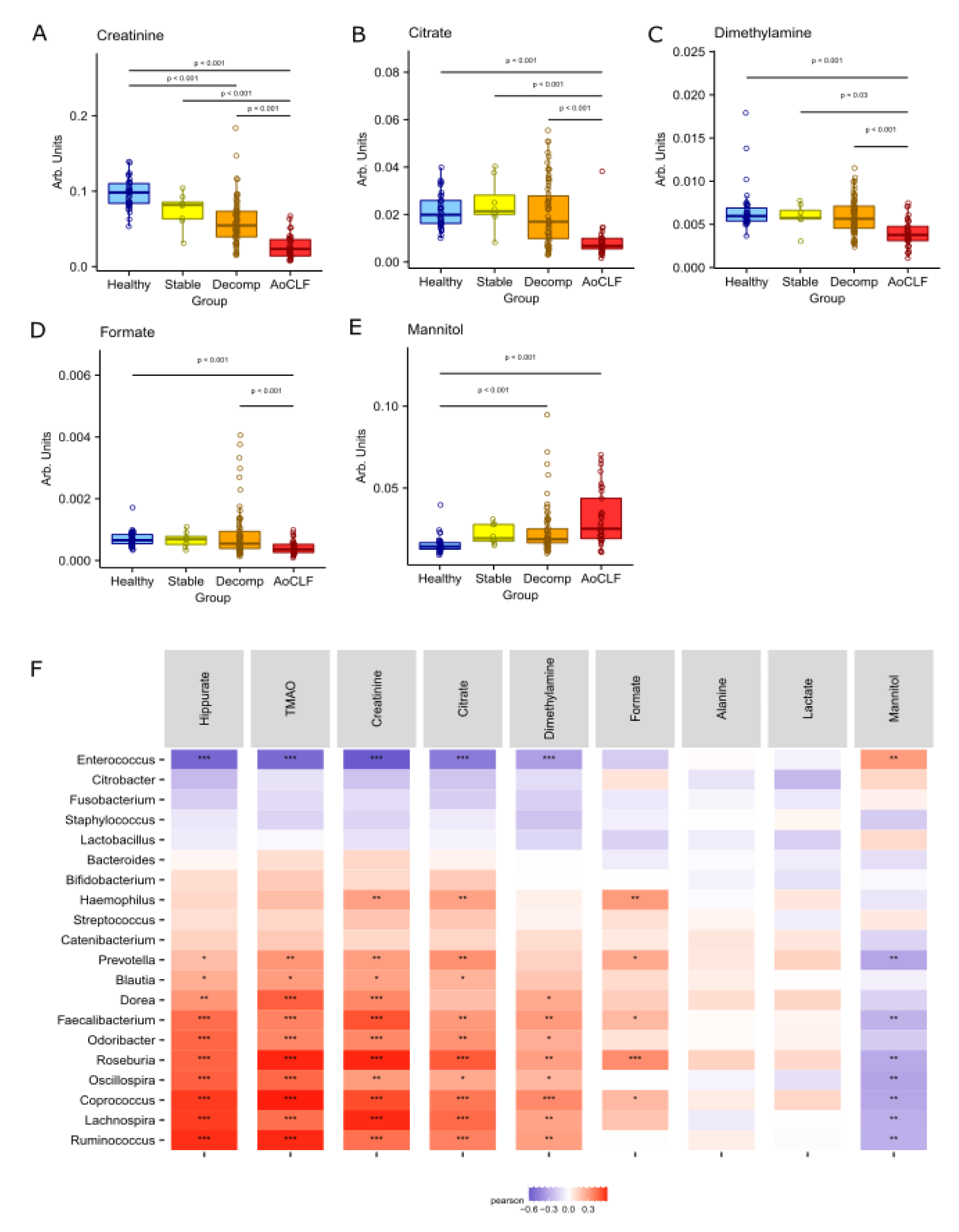
Individual urinary metabolites by cirrhosis severity and correlations between genus proportion and plasma metabolites. (A) Creatinine, (B) Citrate, (C) Dimethylamine, (D) Formate, (E) Mannitol, (F) Correlations between normalised urinary metabolites and normalised genus relative abundance using a Pearson correlation. P-value: * = p<0.05, ** = p<0.01, ** = p<0.001. Significance corrected for multiple testing.

**Supplementary Figure 5.**
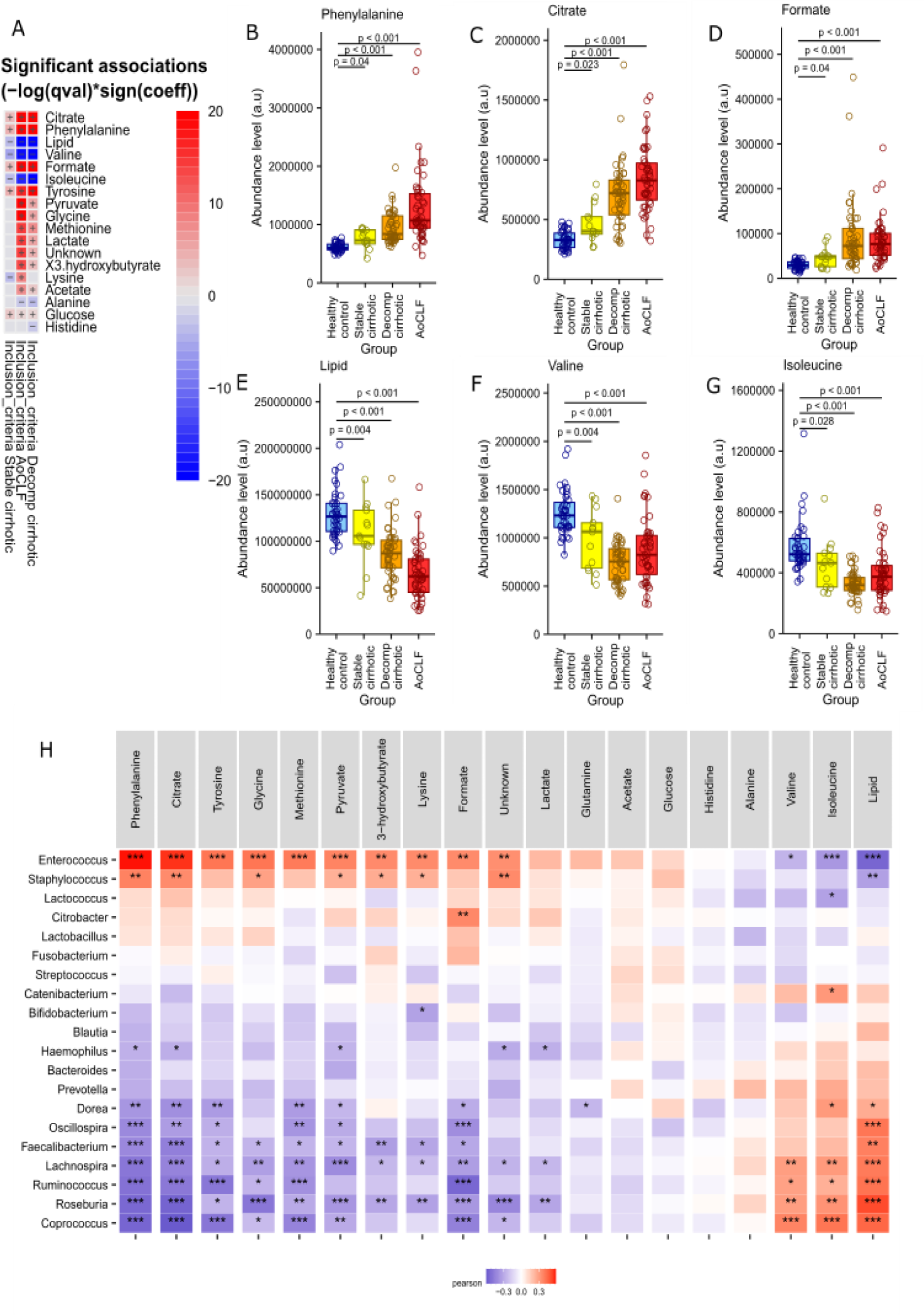
Individual plasma metabolites by cirrhosis severity and correlations between genus proportion and plasma metabolites. (A) Phenylalanine, (B) Citrate, (C) Formate, (D) Lipid, (E) Valine, (F) Isoleucine, (G) Correlations between normalised plasma metabolites and normalised genus relative abundance using a Pearson correlation. P-value: * = p<0.05, ** = p<0.01, *** = p<0.001. Significance corrected for multiple testing.

**Supplementary Figure 6.**
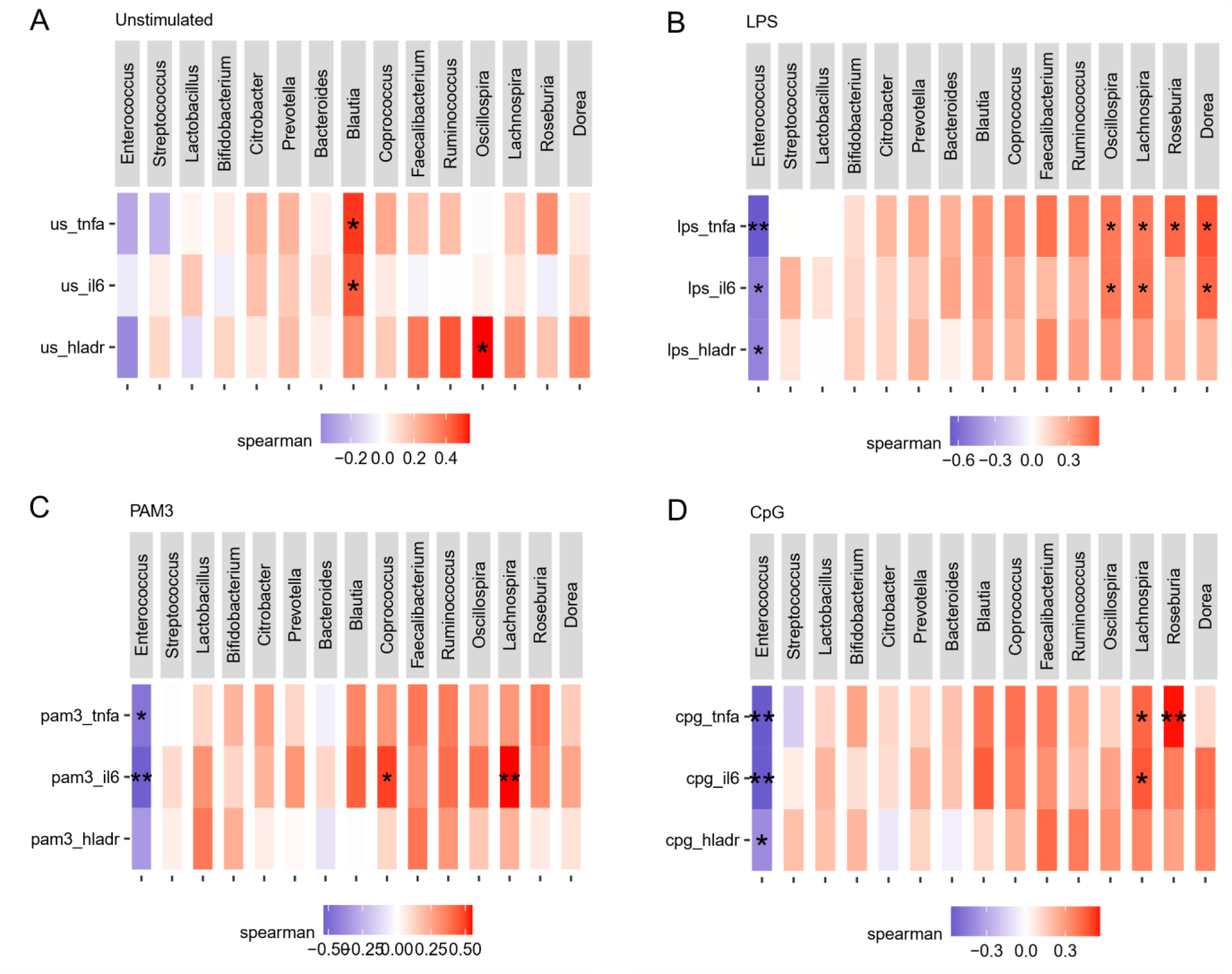
Correlations between genus proportion and monocyte stimulation. Full correlations between the top 15 most abundant genera and TNF-alpha, IL-6, and HLA-DR percentage in (A) Unstimulated, (B) LPS stimulated, (C) PAM3 stimulated, and (D) CpG simulated monocytes. Correlations between normalised plasma metabolites and normalised genus relative abundance using a Pearson correlation. P-value: * = p<0.05, ** = p<0.01. Significance corrected for multiple testing.

**Supplementary Figure 7.**
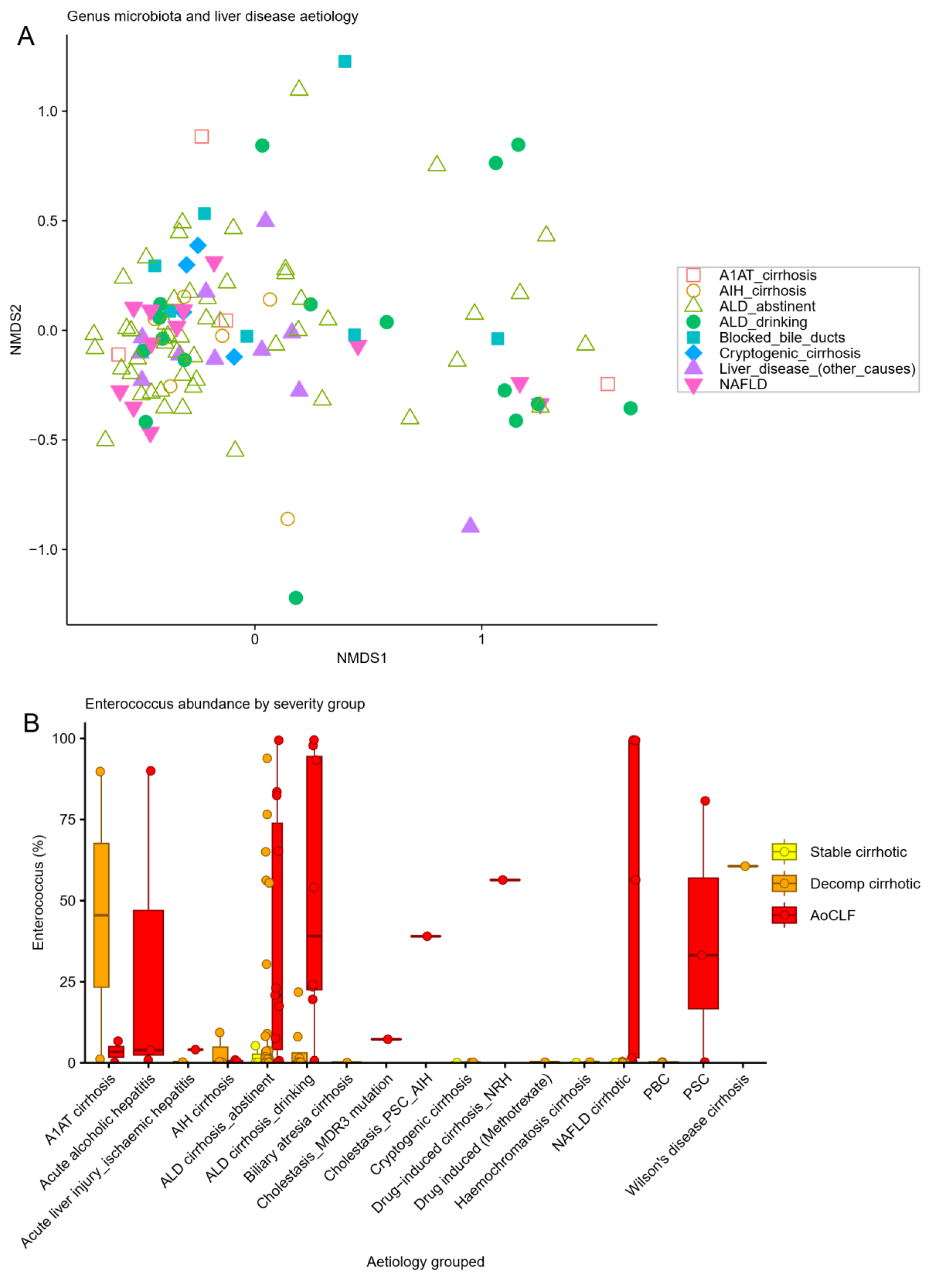
Aetiology associations with overall microbiota composition and *Enterococcus* abundance. (A) NMDS clustering of overall genus level faecal microbiota composition based on a Bray-Curtis matrix showing patient samples with patient aetiology condensed to eight groups for the purposes of analysis showing a lack of clustering by aetiology (PERMANOVA: P = 0.1). (B) Box plots show full patient aetiology groups with faecal *Enterococcus* abundance grouped by illness severity.

## Notes

### Author Declarations

The observational study was granted approval by the NHS Health Research Authority NRES Committee London Westminster [REC reference: 12/LO/1417] and local sponsor Research and Development department (KCH12-126). Ethical approval for the interventional trial was obtained from NHS Health Research Authority NRES Committee South Central-Oxford C (Bristol) [REC reference: 14/SC/0088] and from the Medicines and Healthcare products Regulatory Agency for Clinical Trial Authorisation [EudraCT number: 2013-004708-20; ClinicalTrials.gov NCT02019784]. The studies were conducted in compliance with the principles of the Declaration of Helsinki (1996), principles of Good Clinical Practice, Research Governance Framework and where relevant, the Medicines for Human Use (Clinical Trial) Regulations.

### Summary of Updates

We have removed an author and author order has changed.

